# Changes in neuronal genes in prenatally alcohol-exposed placentas associate with neuropsychological traits at the age of six

**DOI:** 10.64898/2026.01.21.26344516

**Authors:** K Rämö, E Wallén, E Saure, M Jolma, P Auvinen, H Kahila, N Kaminen-Ahola

## Abstract

Prenatal alcohol exposure (PAE) disrupts embryonic development and gives rise to a variable fetal alcohol spectrum disorder (FASD) phenotype characterized by neurodevelopmental and dysmorphological defects. We investigated the effects of PAE on placental gene regulation by performing genome-wide DNA methylation (DNAm) microarray and gene expression (3’mRNA sequencing) analyses in 87 PAE, 77 unexposed control, and 11 only smoking-exposed placentas. Significant alterations were identified in genes involved in synaptic function including both excitatory and inhibitory neurotransmission, and in genes previously associated with attention-deficit/hyperactivity disorder (ADHD), autism spectrum disorder, schizophrenia, and addiction. When placental molecular alterations were compared with neuropsychological and dysmorphological phenotypes of the same children assessed at the age of six, numerous associations were observed for DNAm and gene expression with head circumference as well as cognitive performance, ADHD, and dysmorphology scores. Among the phenotype-associated genes were *NR2F1, GPHN, GLI3,* and *FRZB*, which are involved in neurogenesis and cortical patterning, postsynaptic clustering of GABAα receptors, brain and craniofacial development, and regulation of Wnt signaling, respectively. As these alterations were detected in the placenta, this tissue not only enables the identification of phenotype-specific FASD candidate genes but may also provide a unique window into the developmental mechanisms underlying complex neurodevelopmental and dysmorphological disorders.

## INTRODUCTION

The intrauterine environment has an impact on the development and lifelong health of the individual. Prenatal alcohol exposure (PAE) is one of the most harmful environmental factors and a major public health concern globally^1^. PAE has been associated with a variety of adverse developmental outcomes, including central nervous system (CNS) dysfunction, craniofacial malformations, birth defects, and growth restriction, which fall under the umbrella term of fetal alcohol spectrum disorder (FASD)^2^. The features of complex FASD phenotype overlap with other neurodevelopmental disorders (NDDs)^3,4^, such as learning and memory deficits^5^, attention-deficit/hyperactivity disorder (ADHD), autism spectrum disorder (ASD), as well as with schizophrenia (SCZ), which has a strong neurodevelopmental component^6^. Furthermore, PAE significantly increases the risk of mental health disorders such as depression, anxiety, and addiction^7^.

Several *in vitro* models and animal studies have shown that alcohol exposure during the first developmental weeks is capable of altering epigenetic marks, such as DNA methylation (DNAm), affecting gene expression, and disrupting embryonic developmental programming^8–10^. The developing nervous system is particularly sensitive to the effects of PAE^11,12^. Among the gastrulating human germ layer cells, the DNAm and gene expression of ectodermal cells are especially susceptible to the effects of *in vitro* alcohol exposure^12^. Furthermore, substantial PAE limited within the first eight gestational weeks (GWs) is sufficient to result in FASD with clinically significant neurocognitive impairment^13^. Indeed, alcohol-related neurodevelopmental disorders (ARNDs) constitute the most common subgroup of FASD^14^. It has been shown that alcohol disrupts synaptic function and plasticity subsequently affecting the formation and remodeling of neuronal networks^15^. Consistent with this, altered synaptic and mitochondrial gene expression, an imbalance of excitatory/inhibitory (E/I) synaptic activity in the brain, and cognitive and behavioral abnormalities have been observed in a PAE mouse model^16^. In humans, altered E/I balance has previously been linked to NDDs, including cognitive disorders, as well as neuropsychiatric conditions^17–20^.

The placenta is a temporary organ that not only transfers nutrients, oxygen, and antibodies from the maternal circulation to the fetus but also produces growth hormones and neuropeptides, and acts as a partial barrier against harmful substances^21^. It is an easily accessible tissue, and recent studies have suggested that it could be a useful tissue to investigate the effects of the intrauterine environment on human development. Differentially methylated regions (DMRs) in the placenta have been shown to be enriched in genes involved in neuronal development and synaptic function^22–24^. Also, similar NDD-associated DNAm profiles and gene expression have been observed in both placenta and brain tissues in human^24^ and mouse^25^. Some previously observed associations between placental DNAm and later neurodevelopmental outcomes, including cognitive function and ASD in human^26,27^, suggest that the placenta may offer molecular insights into NDDs and provide early epigenetic markers of neurodevelopmental risk. The mechanisms that underlie the observed associations remain obscure, but recent findings have heightened interest in the poorly understood placenta–brain axis^28^ and neuroplacentology^29^.

To explore the molecular etiology of FASD, we have collected placental samples from prenatally alcohol- and smoking-exposed as well as control newborns. Here, we focused on the effects of PAE on the placental methylome and transcriptome and minimized the effects of smoking, a common confounding factor, by adding smoking-exposed-only (SEO) placentas to the control group. Furthermore, the observed PAE-associated alterations were confirmed by excluding all smoking-exposed placentas and comparing a subgroup of only alcohol-exposed placentas to the controls. To explore potential links between the observed PAE-associated molecular alterations in the placenta and phenotypes, we have performed neuropsychological and dysmorphological assessments on the same children at the age of six^13^. The majority (76%) of the tested PAE children met the criteria of FASD at this age. Finally, we performed association analyses between the DNAm and expression of placental PAE-associated genes and head circumference (HC) as well as cognitive, ADHD, FAS dysmorphology, and midfacial hypoplasia scores of the children.

## RESULTS

### Characteristics of the cohort

General characteristics of 87 PAE and 88 control (including 77 unexposed and 11 SEO) newborns as well as their mothers were compared (Supplementary Table S1). PAE newborns had significantly lower gestational age (*P*=0.001, Mann-Whitney U), lower birth weight (standard deviation, SD, hereafter denoting z-scores) (*P*=0.021, Student’s t-test), lower birth length (SD) (*P*=0.019, Student’s t-test), and smaller birth HC (SD) (*P*=0.003, Student’s t-test) compared to controls and smoking-exposed newborns.

Mothers of PAE newborns had significantly higher pre-pregnancy BMI (*P*=0.017, Mann-Whitney U) and lower gestational weight gain (*P*=0.015, Student’s t-test) compared to control mothers. After exclusion of all smoking-exposed samples, 16 only PAE and 77 unexposed control newborns as well as their mothers were compared (Supplementary Table S1). The mothers of only PAE newborns still had higher pre-pregnancy BMI (*P*<0.001, Mann-Whitney U).

### PAE-associated genome-wide DNAm in placenta

Genome-wide DNAm analysis by microarrays (EPIC, Illumina) for 87 PAE and 88 control (including 77 unexposed and 11 SEO) placental samples resulted in 3862 significantly differentially methylated PAE-associated CpG sites (53% hypomethylated) with FDR<0.05 (Fig. 1a, Supplementary Table S2). To separate the most prominent changes and minimize false positive hits due to the observed inflation, we focused on the CpG sites with DNAm difference in the effect size of ≥5% between PAE and control placentas (FDR<0.05, Δβ≤−0.05 and Δβ≥0.05), which were termed as differentially methylated positions (DMPs). We observed 225 PAE associating DMPs (22% hypomethylated) linking to 154 genes.

**Figure 1.**
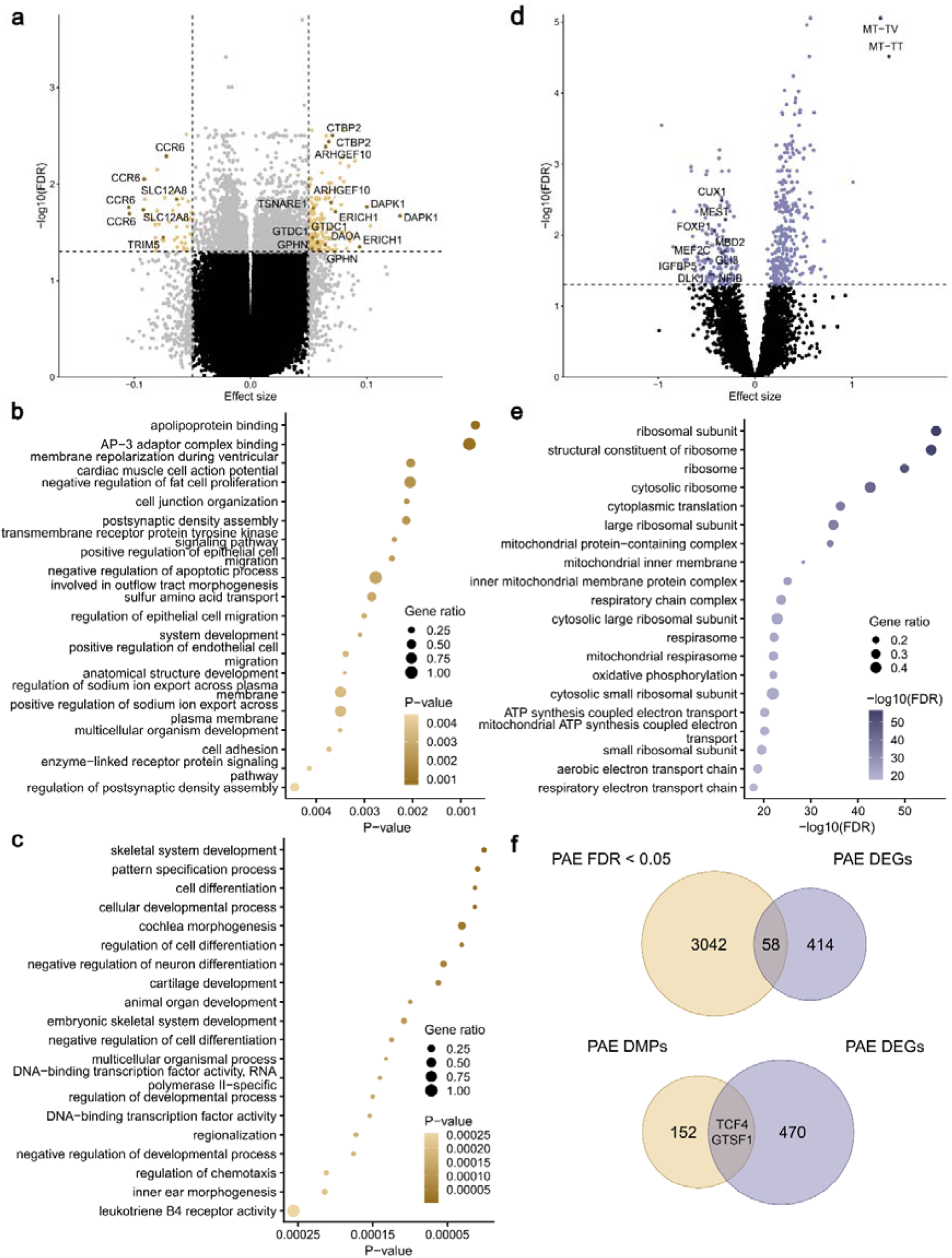
PAE-associated differential DNAm, gene expression, and canonical pathways in the placenta. **a)** Volcano plot showing the distribution of associations between placental CpG sites and PAE. Horizontal line marks FDR 0.05 and vertical line marks effect size ± 0.05. Enriched terms identified in GO pathway analysis of PAE-associated **b)** differentially methylated CpGs (FDR<0.05) and **c)** DMRs in the placenta. In pathway analyses, the 20 most significant pathways are shown. **d)** Volcano plot showing the distribution of associations between mRNA expression and PAE. Horizontal line marks FDR 0.05. **e)** Enriched terms identified in GO pathway analysis of PAE-associated DEGs (FDR<0.05) in the placenta. Venn diagram showing the number of **f)** PAE-associated CpGs (FDR<0.05), DEGs, and the common genes, and PAE-associated DMPs, DEGs, and the common genes.

Several PAE-associated hypomethylated DMPs linked to *CCR6, TRIM5,* and *SLC12A8*, and hypermethylated to *DAPK1, ERICH1*, *ARHGEF10, DAOA, CTBP2, TSNARE1, GTDC1,* and *GPHN* (Fig. 1a). The most hypomethylated DMPs associated with *CCR6*, a key player in immune responses involving inflammation^30^, whereas the most hypermethylated with *DAPK1,* a mediator of gamma-interferon induced programmed cell death, and *ERICH1.* The highest number of hypermethylated DMPs associated with *ARHGEF10* (*rho guanine nucleotide exchange factor 10*), which regulates the activity of small Rho GTPases, proteins crucial for neuronal shape, polarity, migration, and connectivity during brain development^31^.

Analysis of DMRs, defined as regions containing three or more CpGs within a maximum genomic distance of 1000 bp, identified 506 placental DMRs associated with PAE (Supplementary Table S3). Among the genes associated with the most hypomethylated DMRs were *CCR6, GSDMD, GTSF1,* and *HOXC6,* as well as imprinted *FAM50B* and *PEG10.* The most hypermethylated DMRs associated with *ESRRG*, *TSNARE1, SATB2, FRZB*, and *ARHGEF10*. A total of 11 hypermethylated DMRs were associated with *TSNARE1* (*T-SNARE domain containing 1*), which primarily acts as a regulator of endosomal trafficking in neurons, especially in the brain^32^.

According to the Gene Ontology (GO) pathway analysis, all significant alterations in placental DNAm were enriched most significantly in terms such as membrane repolarization during ventricular cardiac muscle cell action potential, negative regulation of fat cell proliferation, cell junction organization, and postsynaptic density assembly (*P*<0.05) (Fig. 1b, Supplementary Table S4). PAE-associated alterations in DMRs enriched in terms such as skeletal system development, negative regulation of neuron differentiation, cartilage development, as well as cochlea and inner ear morphogenesis (*P*<0.05) (Fig. 1c, Supplementary Table S5). Interestingly, all these processes have been previously associated with FASD phenotype^2,33^.

### PAE-associated mRNA expression in placenta

mRNA-seq for 61 PAE and 48 control (including 41 unexposed and 7 SEO) placental samples revealed 472 significantly differentially expressed genes (DEGs) (FDR<0.05) of which 68% were upregulated (Fig. 1d, Supplementary Table S6). The most upregulated genes were MT-TT and MT-TV, which encode mitochondrial tRNA molecules for threonine and valine, respectively. According to the GO enrichment analysis, PAE-associated gene expression is linked predominantly to pathways related to ribosomes as well as mitochondrial protein-containing complex, inner membrane, and respiratory chain complex (FDR<0.05, Fig. 1e, Supplementary Table S7). According to the MitoXplorer database^34^, 19% of DEGs associated with mitochondrial functions, mainly with oxidative phosphorylation, translation, and reactive oxygen species defence (Supplementary Table S8). A total of 58 genes were in common between PAE-associated CpGs (FDR<0.05) and DEGs, and *TCF4* and *GTSF1* were the only genes overlapping between genes associated with DMPs and DEGs (Fig. 1f, Supplementary Table S9). *TCF4* (*transcription factor 4*) is a controller of gene expression in the brain, muscles, and during embryonic development, and it is critical for the development of inhibitory interneurons^35,36^. *GTSF1* (*gametocyte specific factor 1*) is a co-factor in piRNA pathway and is involved in the silencing of transposable elements and protection of genome integrity in germ cells^37^.

### PAE-associated alterations in neurodevelopmental genes

In addition to several developmentally essential PAE-associated genes, such as hypermethylated *HOXA1, MNX1,* and *NODAL,* as well as downregulated *IGFBP5, DLK1, MEF2C, NFIB*, *FOXP1, CUX1, GLI3, MBD2,* and *MEST*, we observed numerous genes involved in synaptic function according to the SynGO database^38^. DMPs, mainly hypermethylated, were associated with 18 synaptic genes (Table 1a). The only hypomethylated DMPs were in *GRIN3A* and *NRN1*, which both associate with presynaptic modulation of chemical synaptic transmission. Moreover, a total of 73 synapse-associated DEGs, including 42 upregulated ribosomal genes (Supplementary Table S10) and 31 other genes (48% upregulated) (Table 1b), were observed.

**Table 1.**
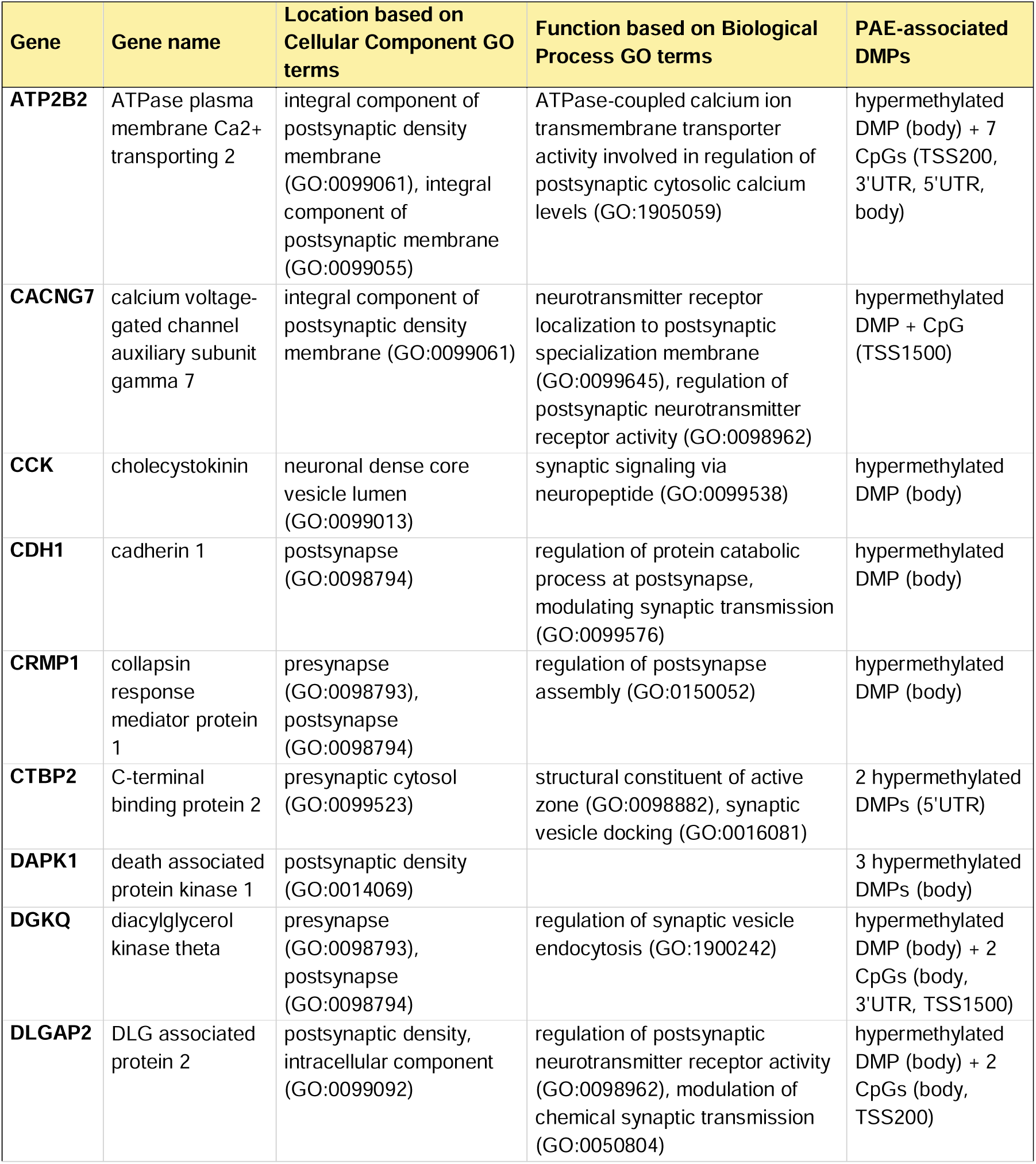
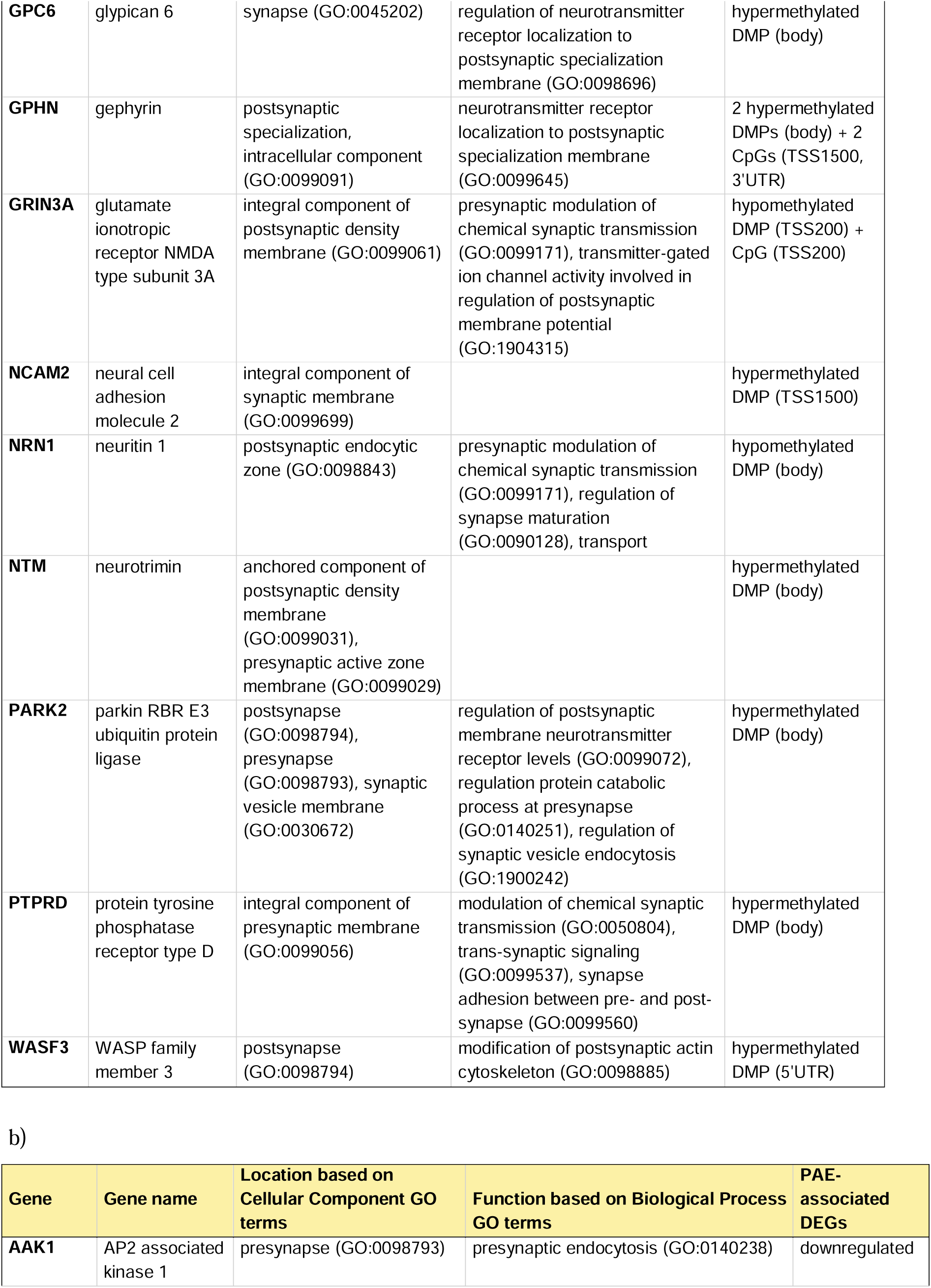
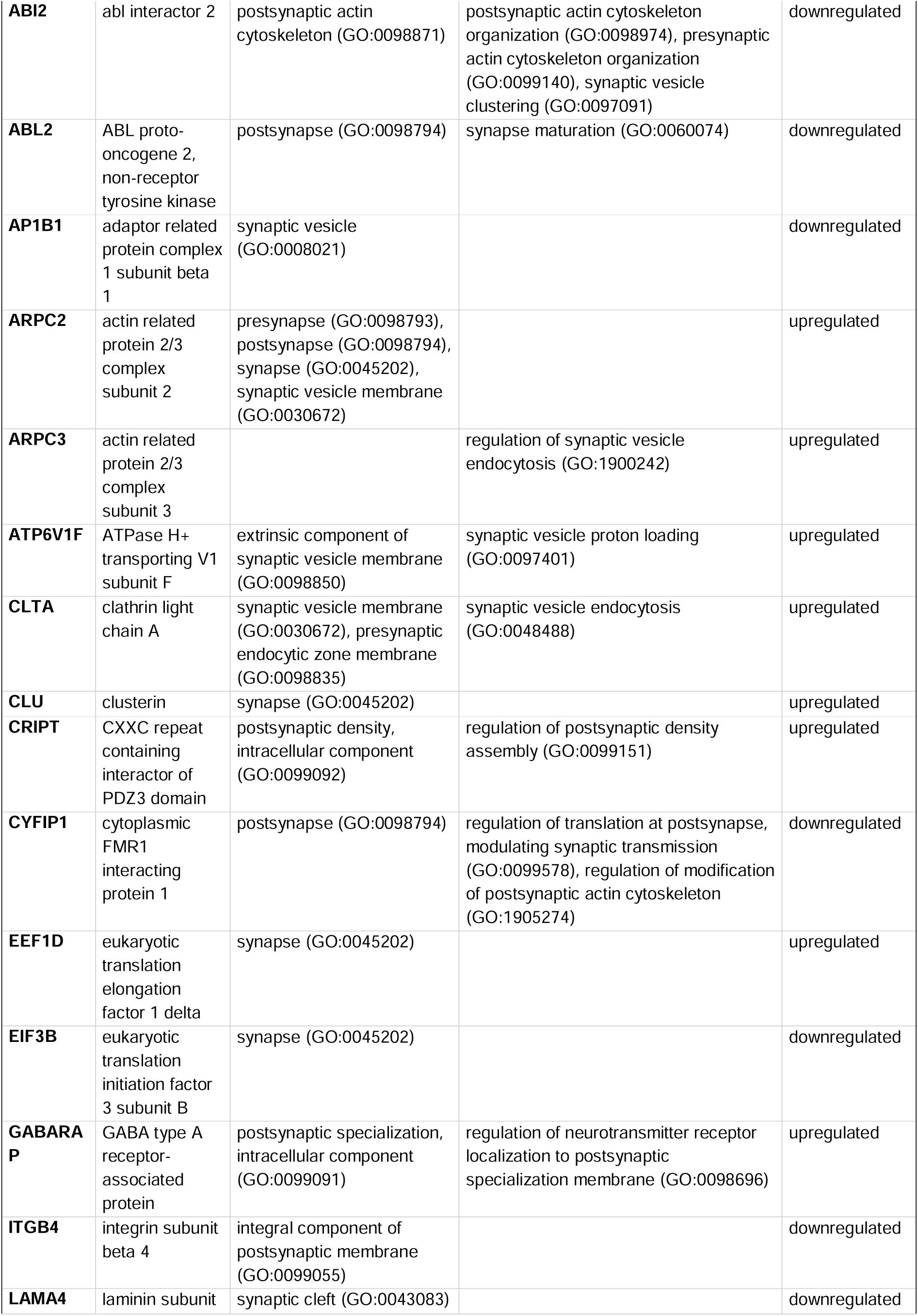
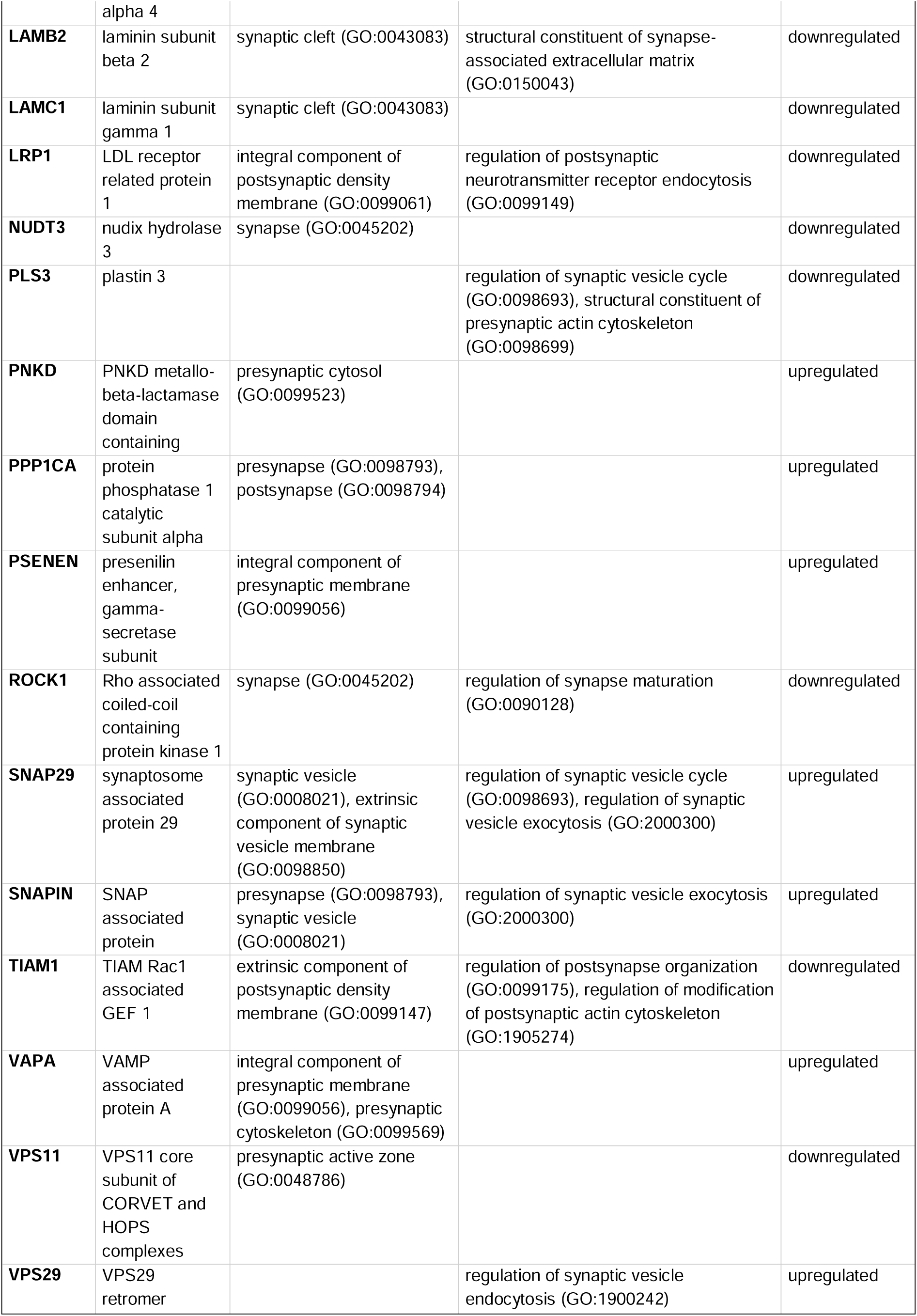
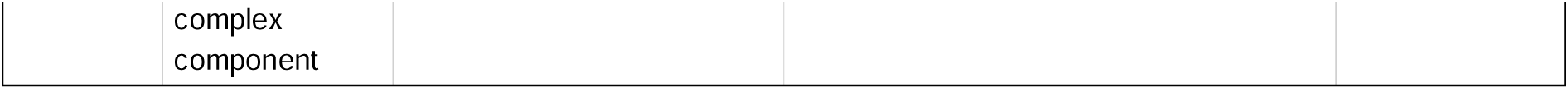
Synaptic genes linked to PAE-associated alterations in DNAm (DMPs) and gene expression (DEGs) in the placenta. Genes, their cellular component locations, and involved biological processes based on **a)** DNAm analysis and **b)** gene expression analysis according to SynGO database. DMP-associated CpGs (FDR<0.05) have also been reported.

Furthermore, in this study, 52 (8.3%) PAE-associated genes have been previously associated with ADHD^39^, 49 (7.6%) with ASD^40^, and 218 (34.9%) with SCZ^41^ (Fig. 2a-c, Supplementary Table S11). A total of 250 genes has been linked to all three disorders across the used resources (Fig. 2d), and 16 of these genes overlap with PAE-associated genes identified in this study (Fig. 2e, Supplementary Table S11). The majority of the 16 genes are involved in embryonic brain development, neuronal differentiation, synapse formation, and epigenetic regulation. Additionally, many of the altered genes observed in this study, such as *ARHGEF10*^42^*, DLGAP2*^43^*, FOS*^44^*, GATA4*^45^, *GPHN*^46^*, GRIN3A*^47^, and *TACR3*^48^, have previously been associated with alcohol addiction or dependence.

**Figure 2.**
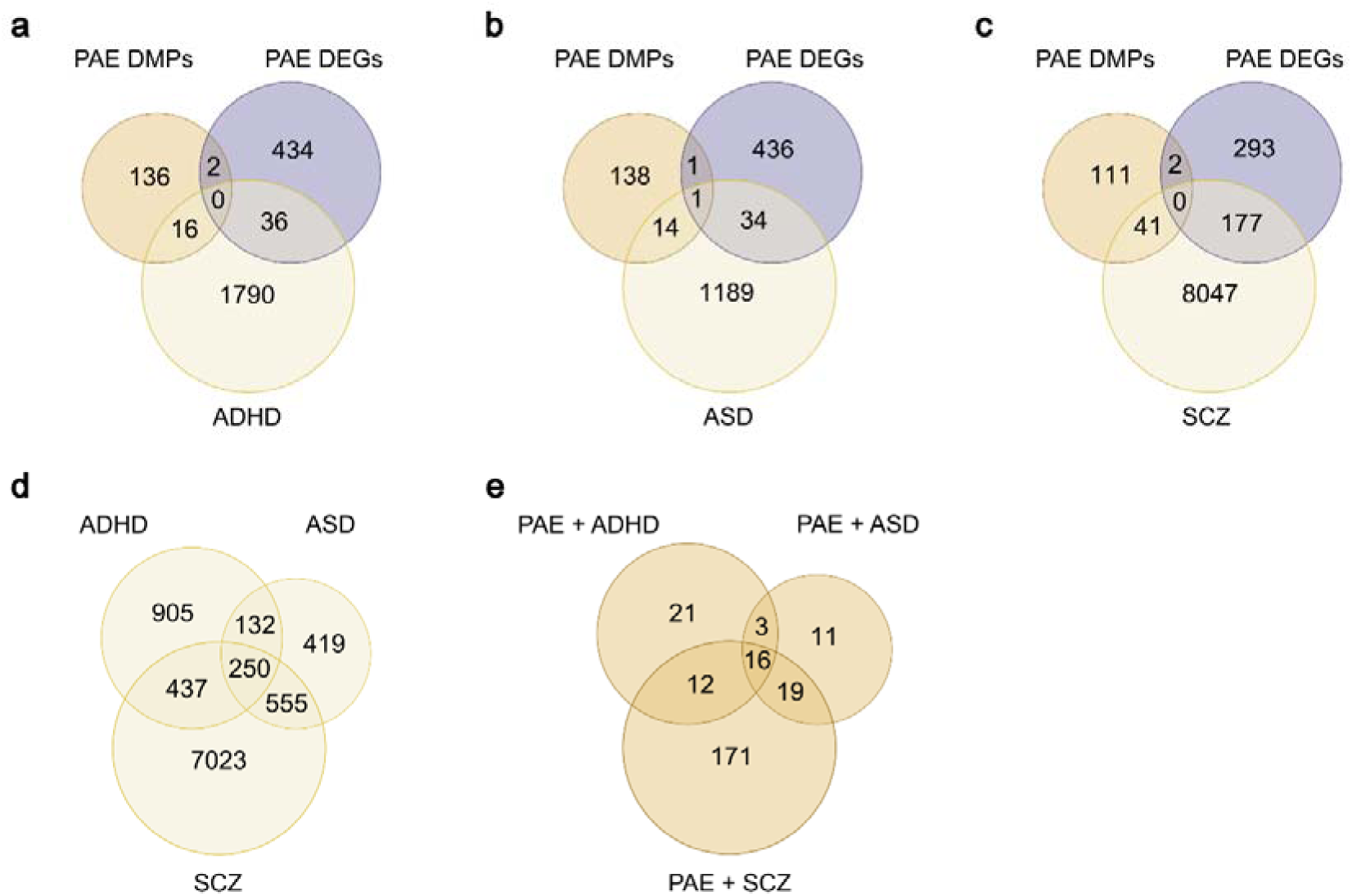
Number of genes identified by PAE-associated DNAm and gene expression analyses that overlap with NDDs sharing similar phenotypic features. PAE-associated DMPs and DEGs were compared to genes previously associated with **a)** ADHD, **b)** autism spectrum disorder (ASD), and **c)** schizophrenia (SCZ) (Hongyao et al. 2023, SFARI, and SZDB, respectively). Venn diagrams showing **d)** overlap with ADHD-, ASD-, and SCZ-associated genes in previous studies, as well as **e)** overlap with ADHD-, ASD-, and SCZ-linked genes that are associated with PAE in the current study, are also presented.

### Sensitivity tests for DNAm and gene expression analyses

The PAE-specificity of the observed alterations was confirmed by performing genome-wide analysis without smoking-exposed samples. In the DNAm analysis (only PAE *n*=16 and controls *n*=77), we observed 108 significantly differentially methylated CpG sites with FDR<0.05, of which 30% were hypomethylated (Supplementary Fig. S1, Supplementary Table S12). A total of five genes were involved in synaptic function according to the SynGO database^38^ (Supplementary Table S10). The highest number of PAE-associated hypermethylated probes were linked to *GPHN* (*gephyrin*), a protein involved in the postsynaptic clustering of GABA alpha and glycine receptors^49^. Among the 55 DMPs, the most hypomethylated one was associated with a neurokinin B signaling mediator *TACR3*. The same gene was also the most hypomethylated gene in our previous study, in which SEO placentas were not included in the controls^23^.

In the gene expression sensitivity analysis (only PAE *n*=10 and controls *n*=41), we observed 295 DEGs (FDR<0.05) of which 77% were upregulated (Supplementary Fig. S1, Supplementary Table S13). Consistently with the main mRNA-seq analysis, PAE-associated alterations were enriched most significantly with ribosomes and mitochondrial functions (Supplementary Table S7). Approximately 50% of the previously observed synapse-specific DEGs were detected in this comparison, such as upregulated *GABARAP* (*GABA type A receptor-associated protein*), involved in GABA receptor trafficking^50^.

### Placental DNAm and gene expression associated with children’s phenotype

In our recent study, we performed neuropsychological, neurodevelopmental, and dysmorphological assessments for 28 PAE and 52 unexposed six-year-old children^13^, whose placentas we have collected at birth. Smaller HCs, lower cognitive performance scores, higher ADHD symptom scores, and dysmorphological features such as midfacial hypoplasia were significantly associated with PAE. Furthermore, 76% of the tested PAE children met the diagnostic criteria for FASD at this age.

In the current study, to explore potential associations between observed molecular placental alterations and phenotypic features, the phenotypic and DNAm data from a total of 83 (30 PAE, 48 control, and five SEO) and mRNA-seq data from 49 (22 PAE, 25 control, and two SEO) placentas of the same six-year-olds were used (Supplementary Table S14, Supplementary Table S15). We performed association analyses comparing DMPs and DEGs to HCs (SD) as well as cognitive, ADHD Rating Scale (ADHD-RS), FAS dysmorphology, and midfacial hypoplasia scores. DMPs and DEGs identified in either of the main or sensitivity analyses, as well as differentially methylated CpGs (FDR<0.05) associated with the same genes as the DMPs, were used in the analyses. Several associations with nominally significant *P*-values were observed (Supplementary Table S16).

#### Placental alterations associated with HC at birth and at the age of six

Since HC is commonly used as an indirect marker of early brain development^51^ and smaller HCs (SD) associated with PAE (Fig. 3a, Supplementary Table S1, Supplementary Table S14), we first analyzed associations between DNAm, gene expression, and HC (SD) at birth and at the age of six (Fig. 3b, Supplementary Table S16). At birth, DNAm of 15 genes were associated with HC (SD), among which *CAMTA1*^52^*, HOXA1*^53^*, MYT1L*^54^*, NR2F1*^55^, and *PTPRD*^56^ have all been associated with HC and/or brain growth also in previous studies (*P*<0.05). Furthermore, a total of 47 correlations was observed between gene expression and HC (SD), of which *BCAP31*^57^, *KMT2C*^58^, *POGZ*^59^, and *RPS6*^60^ have previously been linked with HC and/or brain growth (*P*<0.05, Spearman).

**Figure 3.**
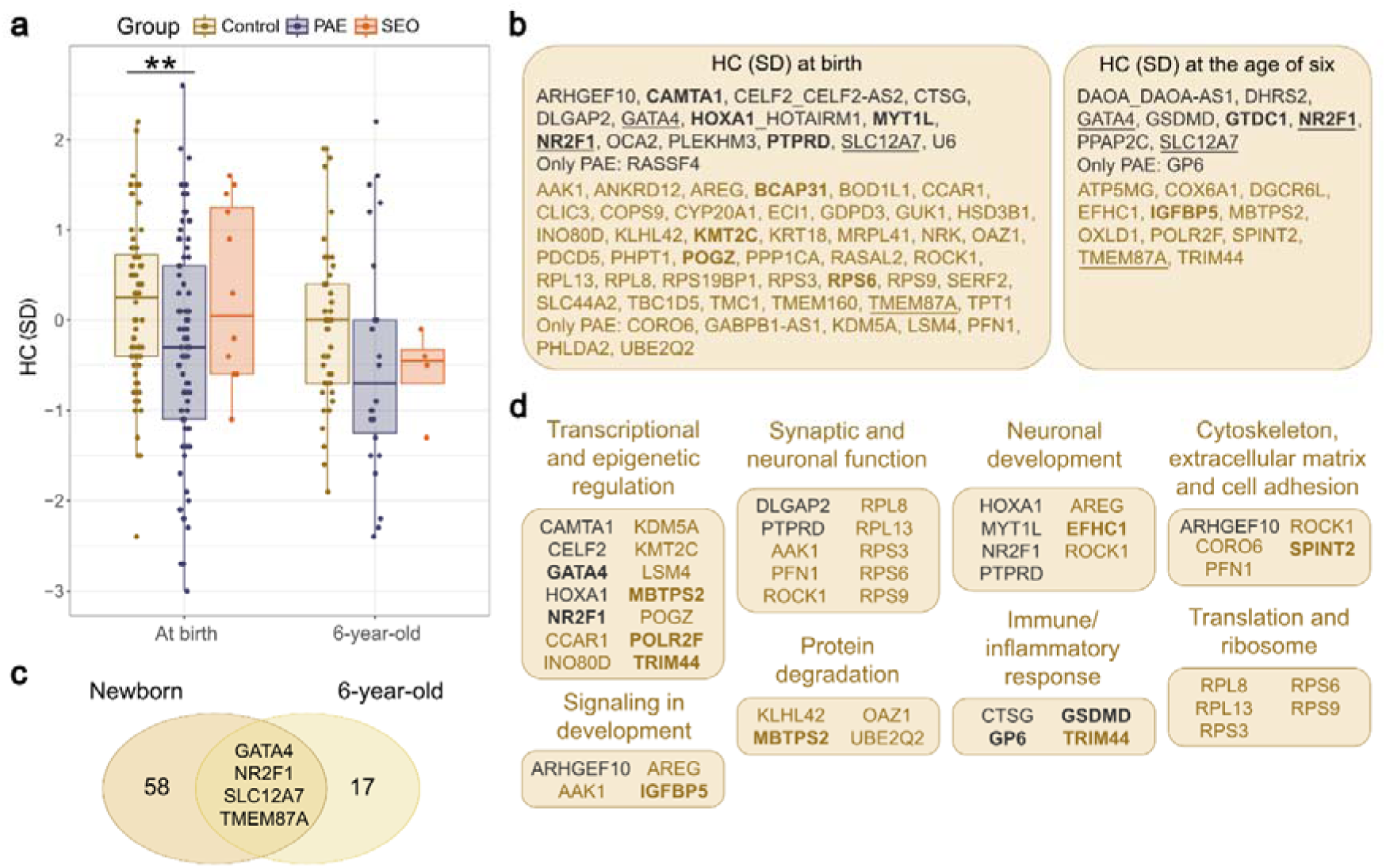
PAE-associated placental alterations associating with HC at birth and at the age of six. **a)** HCs (SD) of PAE newborns and six-year-olds were smaller compared to controls (87 PAE, 76 control, and 12 SEO newborns and 26 PAE, 45 control, and four SEO six-year-olds). **b)** Genes associating with HCs (SD) of newborns (DNAm: *n*=174, gene expression: *n*=109) and at the age of six (DNAm: *n*=75, gene expression: *n*=42) based on DNAm (*P*<0.05) and gene expression (*P*<0.05, Spearman) are shown. Genes previously associated with HC and/or brain growth are shown in bold, and genes associating with HC (SD) at both ages are underlined. Associations between newborns’ HC (SD), DNAm, and gene expression were calculated using data available from the entire cohort. **c)** Common HC (SD)-associated genes between newborns and six-year-olds. **d)** Functional grouping of HC (SD)-associated genes (*P*<0.05) using AI and GO terms. Genes associating with HC at the age of six are shown in bold. Genes associated significantly with HC (SD) in DNAm analyses are in black and in gene expression analyses are in brown (in **b** and **d**). ** *P*-value <0.01, Student’s t-test.

Among 20 genes associating with HC (SD) at six years of age, *GTDC1*^61^, *NR2F1*^55^, and *IGFBP5*^62^ have been previously linked to HC and/or brain growth. Overall, DNAm of transcription factors (TFs) *GATA4* and *NR2F1*, and the potassium–chloride cotransporter *SLC12A7*, as well as expression of the transmembrane protein *TMEM87A*, were associated with HC (SD) both at birth and at the age of six (Fig. 3c). Interestingly, *NR2F1* (*nuclear receptor subfamily 2 group F member 1*) regulates neurogenesis and cortical development, and its pathogenic variants have been associated with neurodevelopmental disorders and brain structural abnormalities^63^.

When HC (SD)-associated genes were grouped according to their function using artificial intelligence (AI) and GO terms, the largest groups were transcriptional and epigenetic regulation, synaptic and neuronal function, neuronal development, as well as cytoskeleton, extracellular matrix and cell adhesion (Fig. 3d, Supplementary Table S17). Changes in gene expression were associated prominently with translation and ribosome as well as protein degradation. Only five terms emerged in the pathway analysis (GO), with ribosome-related terms showing the strongest enrichment (Supplementary Table S18).

#### Placental alterations associated with cognitive performance scores

The cognitive performance scores of Full-Scale Intelligence Quotient (FSIQ) for overall cognitive capacity, Verbal IQ (VIQ) for verbal intellectual abilities, Performance IQ (PIQ) for nonverbal skills, Working Memory Index (WMI) for assessing ability to hold and manipulate information in short-term immediate memory, Processing Speed Quotient (PSQ) for speed and accuracy of processing, and General Language Composite (GLC) for language development were assessed^13^. Significantly lower FSIQ, VIQ, PIQ, and WMI performance scores were observed in PAE children when compared to controls (*P*<0.01, *P*<0.05, *P*<0.01, and *P*<0.01, respectively) (Fig. 4a, Supplementary Table S15). Association analyses between DNAm, gene expression, and cognitive performance scores revealed several genes, which have been previously associated with cognitive impairment and/or intellectual disability, such as *ATP2B2* (associated with PSQ scores in this study)^64^, *CELF2* (WMI)^65^, *CNTN4* (WMI)^66^, *GPHN* (PSQ)^67^, *GTDC1* (PSQ)^61^, and *PTPRD* (PSQ)^68^ in DNAm (*P*<0.05), and *LAMC3* (PIQ)^69^, *LUZP1* (WMI)^70,71^, *MBTPS2* (FSIQ, VIQ, GLC)^72^, *MRPL40* (PSQ)^73,74^, *PSMC3* (WMI)^75^, *RPL10* (FSIQ, VIQ, GLC)^76^, and *UFC1* (FSIQ, PIQ, WMI)^77^ in gene expression (*P*<0.05, Spearman) (Fig. 4b, Supplementary Table S16). Furthermore, two probes of *DAOA* (*D-amino acid oxidase activator*), a modulator at glutamatergic synapses and previously linked to SCZ risk^78^ and verbal fluency^79,80^, were associated with GLC scores (*P*=0.011*, P*=0.012). Also, the expression of *GLI3* (*GLI family zinc finger 3*), which has been previously associated with hippocampus and memory^81^, correlated positively with WMI scores in the current study (*r*=0.31, *P*=0.036, Spearman). The only gene with significant associations with both DNAm (FSIQ, PSQ) and gene expression (FSIQ, VIQ, PIQ) was *CLTA* (*clathrin light chain A*), a component of clathrin coat involved in synaptic vesicles and neurotransmission^82^.

**Figure 4.**
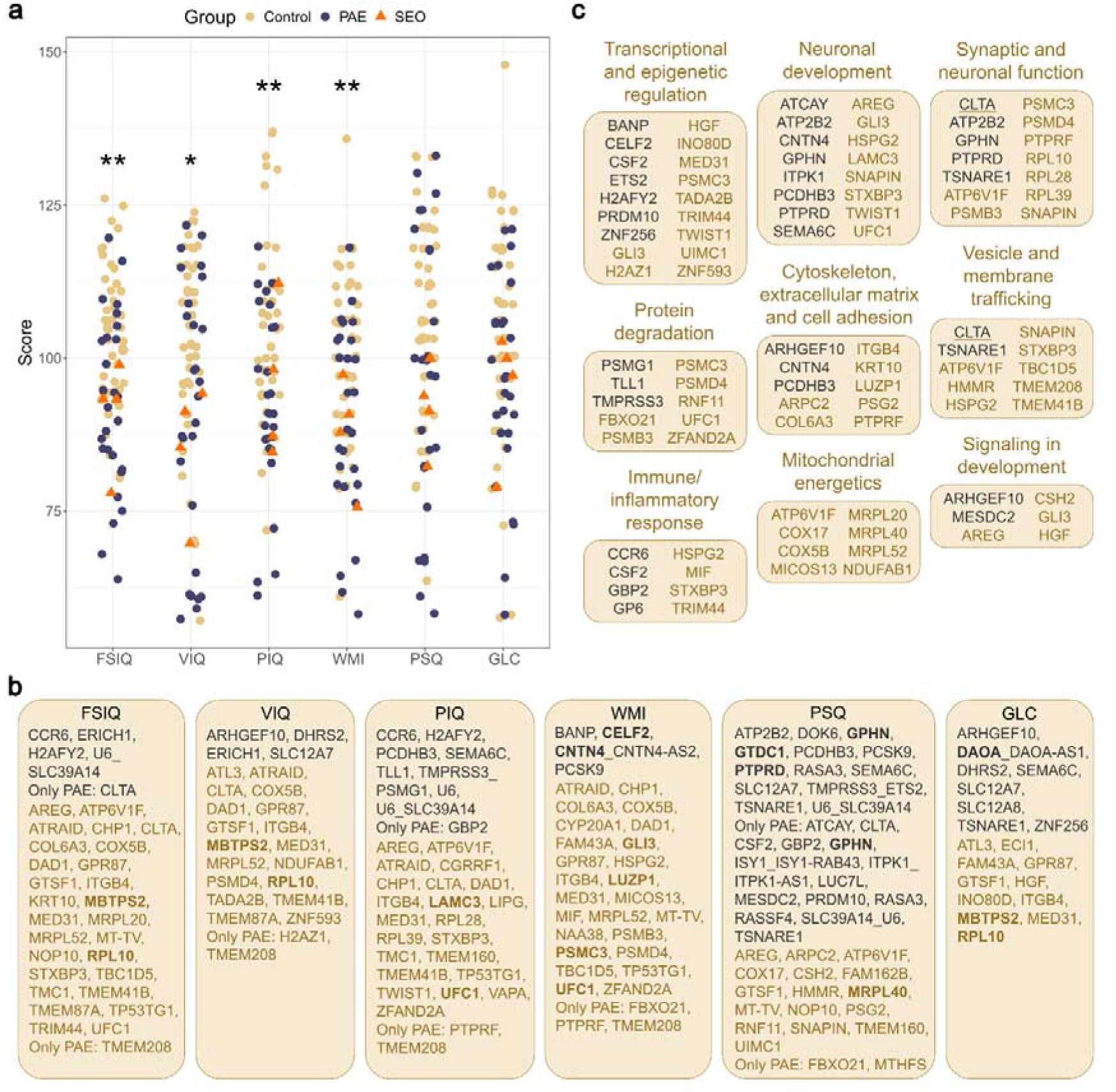
PAE-associated placental alterations and the cognitive performance scores of the same children at the age of six. **a)** Significantly lower FSIQ, VIQ, PIQ, and WMI performance scores were observed in PAE children compared to controls (27 PAE, 48 control, and four SEO six-year-olds). **b)** Genes associating with cognitive performance scores (DNAm: *n*=79, gene expression: *n*=47) based on DNAm (*P*<0.05) and gene expression (*P*<0.05, Spearman) are shown. Genes previously associated with impaired cognition or intellectual disability are shown in bold. **c)** Functional grouping of cognitive performance score-associated genes (*P*<0.05) using AI and GO terms. Underlined genes were significant in both DNAm and gene expression analyses. Genes associated significantly with scores in DNAm analyses are in black and in gene expression analyses are in brown (in **b** and **c**). FSIQ: Full Scale Intelligence Quotient (IQ), VIQ: Verbal IQ, PIQ: Performance IQ, WMI: Working Memory Index, PSQ: Processing Speed Quotient, GLC: General Language Composite. * *P*-value <0.05. ** *P*-value <0.01, Student’s t-test.

When cognitive performance score-associated genes were clustered into functional groups using AI and GO terms, the largest groups were transcriptional and epigenetic regulation, synaptic and neuronal function, neuronal development, as well as mitochondrial energetics including only genes with PAE-associated gene expression (Fig. 4c, Supplementary Table S17). Only five terms emerged in the pathway analysis (GO), with ribosomal terms showing the strongest enrichment (Supplementary Table S18).

#### Placental alterations associated with ADHD-RS scores

ADHD symptom scores, assessed using ADHD-RS questionnaire^13^ completed by parents and teachers, were higher in the PAE group across all domains, including inattention, hyperactivity/impulsivity, and total scores (Fig. 5a, Supplementary Table S15). Among the genes whose DNAm (*P<*0.05) or gene expression (*P*<0.05, Spearman) were associated with ADHD-RS scores, several have also been linked to ADHD genes observed in previous studies (Fig. 5b, Supplementary Table S16). These included *CNTN4*, *CUX1, PARK2*, *POGZ*, and *RAI1* in human^39^, as well as the hyperactivity-associated gene *ARHGEF10* in mouse^83^. Notably, only *CLTA* associated with ADHD-RS scores in both DNAm and gene expression analyses.

**Figure 5.**
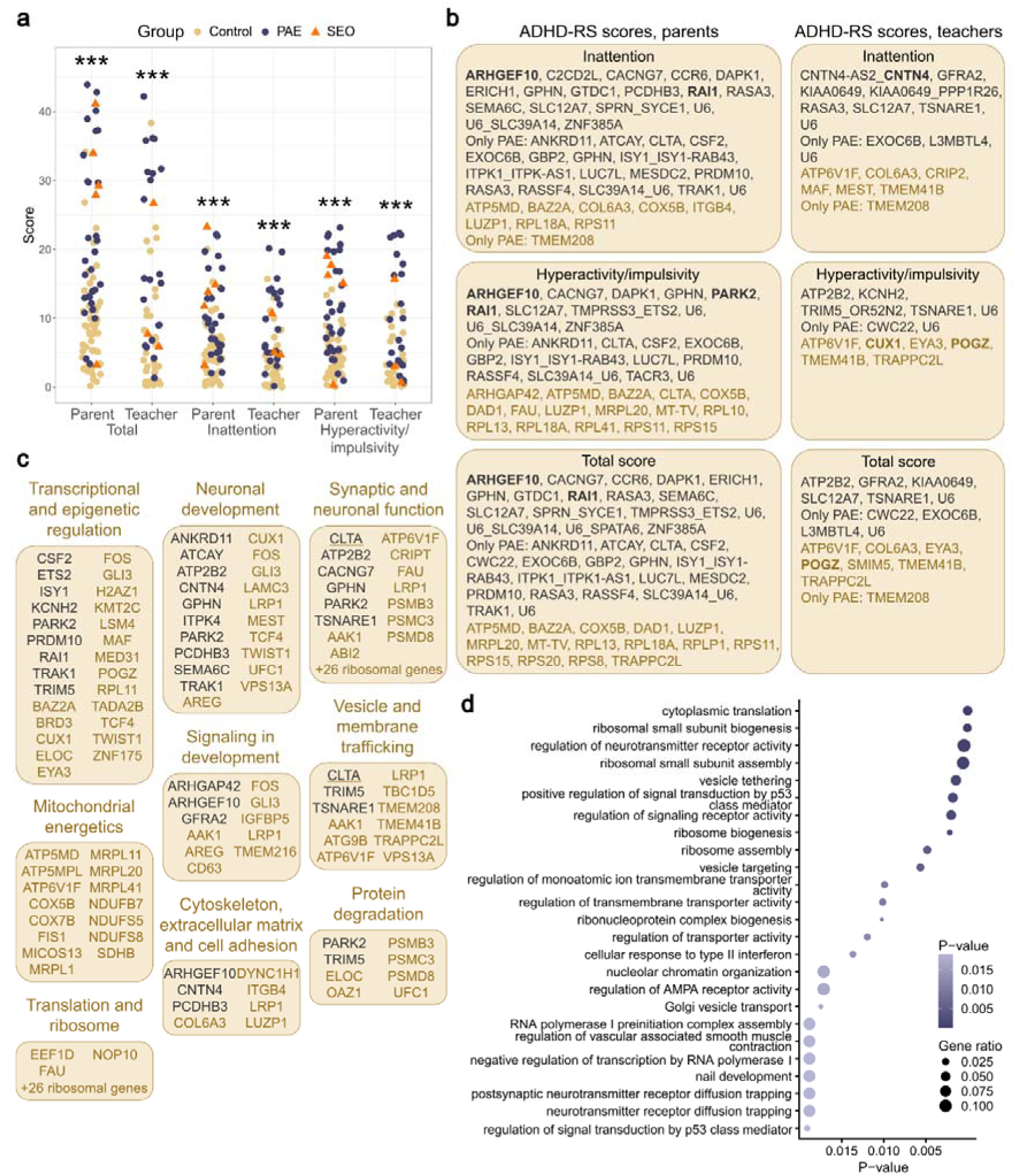
Associations between placental PAE-associated genes and ADHD-RS scores of six-year-old PAE and control children assessed by parents and teachers. **a)** ADHD-RS scores were higher in the PAE group in all categories: inattention, hyperactivity/impulsivity, and total (29 PAE, 48 control, and five SEO six-year-olds). **b)** Genes associating with ADHD-RS scores (DNAm: *n*=82, gene expression: *n*=49 for parents and DNAm: *n*=64, gene expression: *n*=38 for teachers) based on DNAm (*P*<0.05) and gene expression (*P*<0.01, Spearman) are shown. Genes previously associated with ADHD are shown in bold. **c)** Functional grouping of ADHD-RS score-associated genes (*P*<0.05) using AI and GO terms. Underlined genes were significant in both DNAm and gene expression results. **d)** Top 25 most significantly enriched biological process terms identified in GO pathway analysis of ADHD-RS score-associated genes in the placenta. Genes associating significantly with scores in DNAm analyses are in black and in gene expression analyses are in brown (in **b** and **c**). *** *P*-value <0.001, Mann-Whitney U.

When ADHD-RS score-associated genes were grouped according to their function using AI and GO terms, the largest groups were transcriptional and epigenetic regulation, synaptic and neuronal function, neuronal development, vesicle and membrane trafficking, as well as signaling in development (Fig. 5c, Supplementary Table S17). Changes in gene expression were associated prominently with mitochondrial energetics as well as translation and ribosome. According to the pathway analysis (GO), the observed ADHD-RS score-associated genes enriched significantly in addition to ribosomal terms also to terms such as regulation of neurotransmitter receptor activity, vesicle tethering, positive regulation of signal transduction by p53 class mediator, regulation of transmembrane transporter activity, nuclear chromatin organization, regulation of AMPA receptor activity, and postsynaptic neurotransmitter receptor diffusion trapping (Fig. 5d, Supplementary Table S18).

#### Placental alterations associated with FAS dysmorphology scores

Higher FAS dysmorphology scores, including HC (SD), weight/height ratio, height, palpebral fissure length, as well as grades of smooth philtrum and thin vermilion, were associated with PAE^13^ (Fig. 6a, Supplementary Table S14). Among the genes whose placental DNAm (*P*<0.05) or gene expression (*P*<0.05, Spearman) were associated with FAS dysmorphology scores at the age of six, several have also been linked to dysmorphology in previous studies (Fig. 6b, Supplementary Table S16). These included *CEP152*^84^, *DLK1*^85^, *ETS2*^86,87^, *GLI3*^88^, *GTDC1*^61^, *MEST*^89^, *NR2F1*^90^, *RPL10, RPL13A,* and *RPS3*^91^. Functional grouping of FAS dysmorphology score–associated genes using AI and GO terms revealed that, alongside the largest groups—synaptic and neural function, mitochondrial energetics, and transcriptional/epigenetic regulation—a craniofacial/neural crest patterning group was also present (Fig. 6c, Supplementary Table S17). According to the pathway analysis (GO), the observed FAS dysmorphology associated genes enriched most significantly in ribosomal and mitochondrial terms (Supplementary Table S18).

**Figure 6.**
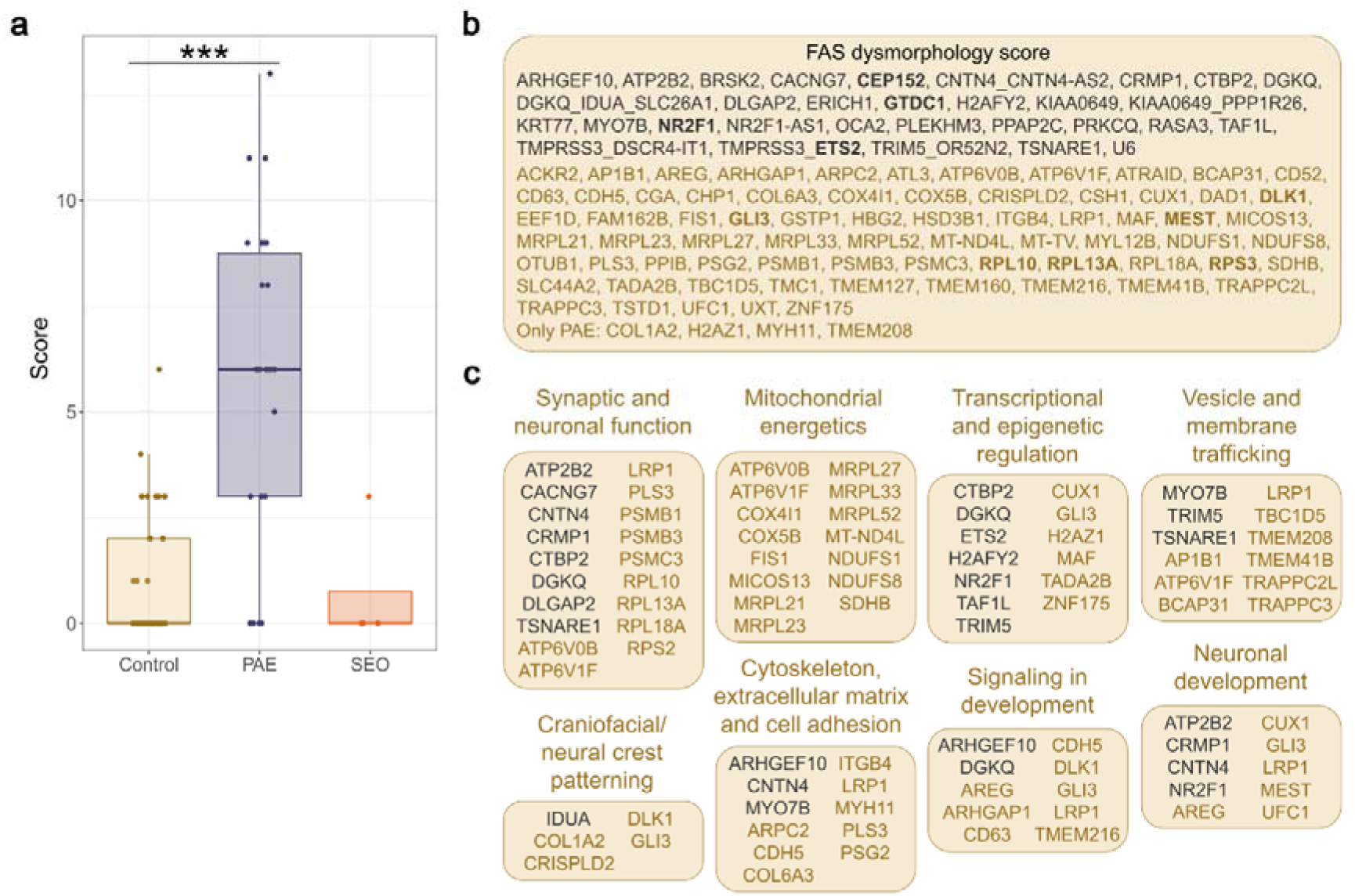
Observed associations between placental PAE-associated genes and FAS dysmorphology scores of six-year-old PAE and control children. **a)** Higher FAS dysmorphology scores were observed in PAE children (26 PAE, 46 control, and four SEO six-year-olds). **b)** Genes associating with FAS dysmorphology (DNAm: *n*=76, gene expression: *n*=43) based on DNAm (*P*<0.05) and gene expression (*P*<0.05, Spearman) are shown. Genes previously associated with dysmorphology are shown in bold. **c)** Functional grouping of FAS dysmorphology-associated genes (*P*<0.05) using AI and GO terms. Genes associating significantly with scores in DNAm analyses are in black and in gene expression analyses are in brown (in **b** and **c**). *** *P*-value <0.001, Mann-Whitney U.

In terms of dysmorphological features, we focused specifically on midfacial hypoplasia, which is not included in the standard FAS facial dysmorphology diagnostic criteria, although it is commonly associated with PAE^92^. Among the ten genes whose placental DNAm was associated with midfacial hypoplasia scores, was hypermethylated *FRZB* (*secreted frizzled-related protein 3*) (*P*=0.05) (Supplementary Table S16), an antagonist of the canonical Wnt signaling pathway^93^. In addition to FAS dysmorphology scores, *GLI3* expression correlated negatively also with midfacial hypoplasia scores (*r*=-0.342, *P*=0.025 and *r*=-0.365, *P*=0.016, respectively). In analyses restricted to the PAE samples, DNAm associated with midfacial hypoplasia at ten genes, including *FRZB* with increased DNAm of two probes (*P* = 0.007 and *P* = 0.04). In the PAE samples, *DKK1* was upregulated, and among the 15 genes whose expression correlated with midfacial hypoplasia scores (*r*=0.496, *P*=0.036, Spearman) (Supplementary Table S16). Interestingly, *DKK1* (*dickkopf WNT signaling pathway inhibitor 1*) encodes a secreted protein that also antagonizes the canonical Wnt/β-catenin signaling, a pathway central to craniofacial patterning^94^. Moreover, its ethanol-induced increased expression and suppressive effect on canonical Wnt/β-catenin signaling in rat cells *in vivo* and *in vitro* has been previously observed^95^.

### Associations between DNAm, gene expression, and phenotype

To elucidate the potential effects of altered DNAm on gene expression, the correlations between DMPs as well as differentially methylated CpGs (FDR<0.05) associated with the same genes as the DMPs at the regulatory regions (TSS1500, TSS200, 5′ UTR and 1stExon) and corresponding gene expression were calculated. This analysis revealed three significantly correlating genes: increased DNAm correlated with downregulated expression of *FRZB,* increased DNAm correlated with increased expression of *CELF2,* and decreased DNAm correlated with upregulated *GTSF1* (Bonferroni corrected *P*<0.05, Pearson) (Supplementary Table S19). Interestingly, the DNAm of the correlating probes of *FRZB* and *GTSF1* were associated with midfacial hypoplasia. Also, the correlations between the expression of DEGs and DNAm of the probes on their corresponding regulatory regions were analyzed. A total of 27 significantly correlating distinct phenotype-associated genes were observed, including *GTSF1* (FSIQ, VIQ, PSQ, GLC)*, DLK1* (FAS dysmorphology)*, LUZP1* (WMI, ADHD), and *LAMC3* (PIQ) (Bonferroni-corrected *P*<0.05, Pearson) (Supplementary Table S19).

### Placental alterations associated with maternal alcohol consumption

Finally, associations between DNAm, gene expression, and self-reported information about maternal alcohol consumption were assessed (Table 2, Supplementary Table S16). Three analyses were performed: (1) maternal AUDIT scores in the full cohort (DNAm *n*=54, gene expression *n*=33), (2) maternal AUDIT scores in the tested six-year-olds (DNAm *n*=18, gene expression *n*=11), and (3) maternal alcohol doses in the full cohort (DNAm *n*=28, gene expression *n*=21). Genes *CCR6*^96^, *GPHN*^46^, *CDH5*^97^*, NUDT3*^98^, *RBM25*^99^, *S100A6*^100^, *VWF*^101^, and *ZFAND2A*^102^, which have previously been associated with alcohol exposure, showed the most consistent associations with maternal alcohol consumption, reaching significance in two of three analyses (FDR<0.05 or *P*<0.05, Spearman). Additionally, other consistently associating genes were *ANKRD11, CLU, CTSO, FAM160B2, MTHFS, NR2F1, OST4, SDHB, SERP1, TMEM59*, and *TRIM44*. Interestingly, *MTHFS* encodes methenyltetrahydrofolate synthetase, an enzyme involved in folate metabolism and the generation of one-carbon units required for nucleotide synthesis and DNAm^103^. Altered methyl donor availability has been proposed as one mechanism underlying the epigenetic effects of PAE^12,104^.

**Table 2.**
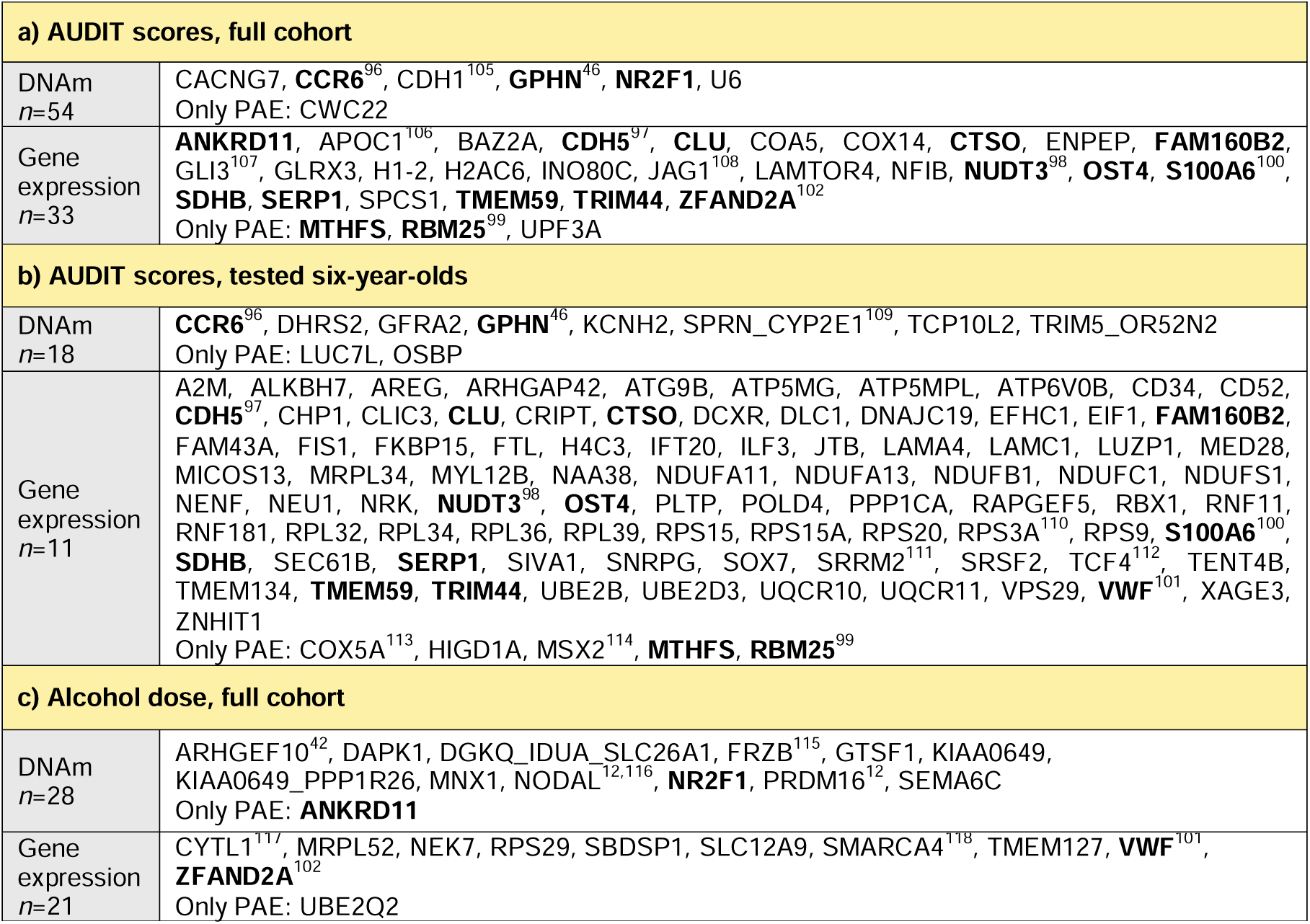
Associations between placental PAE-associated genes and maternal alcohol consumption. Genes, which DNAm (*P*<0.05) and gene expression (*P*<0.05, Spearman) associated with maternal AUDIT scores in the **a)** full cohort, and **b)** tested six-year-olds, as well as with **c)** maternal alcohol doses in the full cohort, are shown. Genes reaching significance in two of three analyses are shown in bold with references included for genes previously associated with alcohol exposure.

## DISCUSSION

To our knowledge, this is the first study to investigate the associations of placental genome-wide DNAm and gene expression with the neuropsychological and dysmorphological characteristics of the same children. By exploring the effects of PAE on placental gene regulation and excluding smoking as a common confounder in PAE studies, we detected a large number of PAE-associated neuronal and mitochondrial genes in the placenta. Surprisingly many of the genes were involved in synaptic function, consistent with the prevailing consensus that alcohol exposure disrupts synaptic balance by affecting E/I transmission^15^. Furthermore, we observed several associations between PAE-associated genes in the placenta and cognitive, ADHD, and dysmorphological traits of the same children at the age of six. Our findings are consistent with the mouse study of Arzua and colleagues, in which they observed numerous PAE-induced changes in synaptic and mitochondrial gene expressions, imbalance of E/I synaptic activity in brain tissue, as well as cognitive and behavioral abnormalities in offspring^16^.

In general, currently observed PAE-associated genes have been identified in several *in vitro* and *in vivo* studies, such as *in vitro* alcohol-exposed gastrulating cells (*ARHGEF10, IGFBP5*, *NODAL, PRDM16, RASA3,* and *SULF1,* several ribosomal genes, as well as imprinted *DLK1*, *MEST,* and *PEG10*)^12^, human cortical brain organoids (in total 48, including *ARHGEF10, CNTN4, FRZB,* and *GRIN3A*)^119^ as well as *in vivo* exposed human buccal epithelial cells (BECs) (*ARHGEF10, ERICH1, NOS1AP,* and *RASA3)*^120,121^, human placenta *(ERICH1* and *DLGAP2)*^121^, human fetal brain (*NR2F1*)^122^, and mouse brain (*LAMA4*)^16^. Also, many of the genes we identified have been previously linked to the same phenotypic traits, supporting our findings. The phenotype-specific associations, together with the limited number of genes correlating with maternal alcohol consumption (AUDIT scores or alcohol doses) indicate that the observed correlations are unlikely to represent merely biomarkers of exposure and may instead reflect biological processes linked to the phenotype itself. Also, PAE-associated altered expression of several ribosomal and mitochondrial genes was observed. This is consistent with previous studies, since alcohol exposure has been associated with impaired ribosome biogenesis^110^ as well as increased oxidative stress and mitochondrial dysfunction in several tissues^123,124^, also in the rat placenta^125^.

### DNAm and gene expression associated with phenotypic traits at the age of six

By comparing observed molecular PAE-associated placental alterations and the phenotypic features of the same children at the age of six, we discovered several new candidate genes for the phenotypic features of FASD, such as *ARHGEF10, CAMTA1, CELF2, CLTA, CTBP2, DAOA, DKK1, DLK1, FRZB, GLI3, GPHN, GTDC1, LAMC3, LUZP1, NR2F1*, *RAI1,* and *TSNARE1*. Many of the genes identified have previously been linked to syndromes with features reminiscent of FASD (Table 3). Compared to DNA sequence mutations, which have mainly been observed in genes responsible for known syndromes, epigenetic changes may indicate subtler alterations in gene function and therefore milder phenotypes. Conversely, studying changes in gene regulation enable to decipher developmentally crucial genes, many of which are rarely mutated.

**Table 3.**
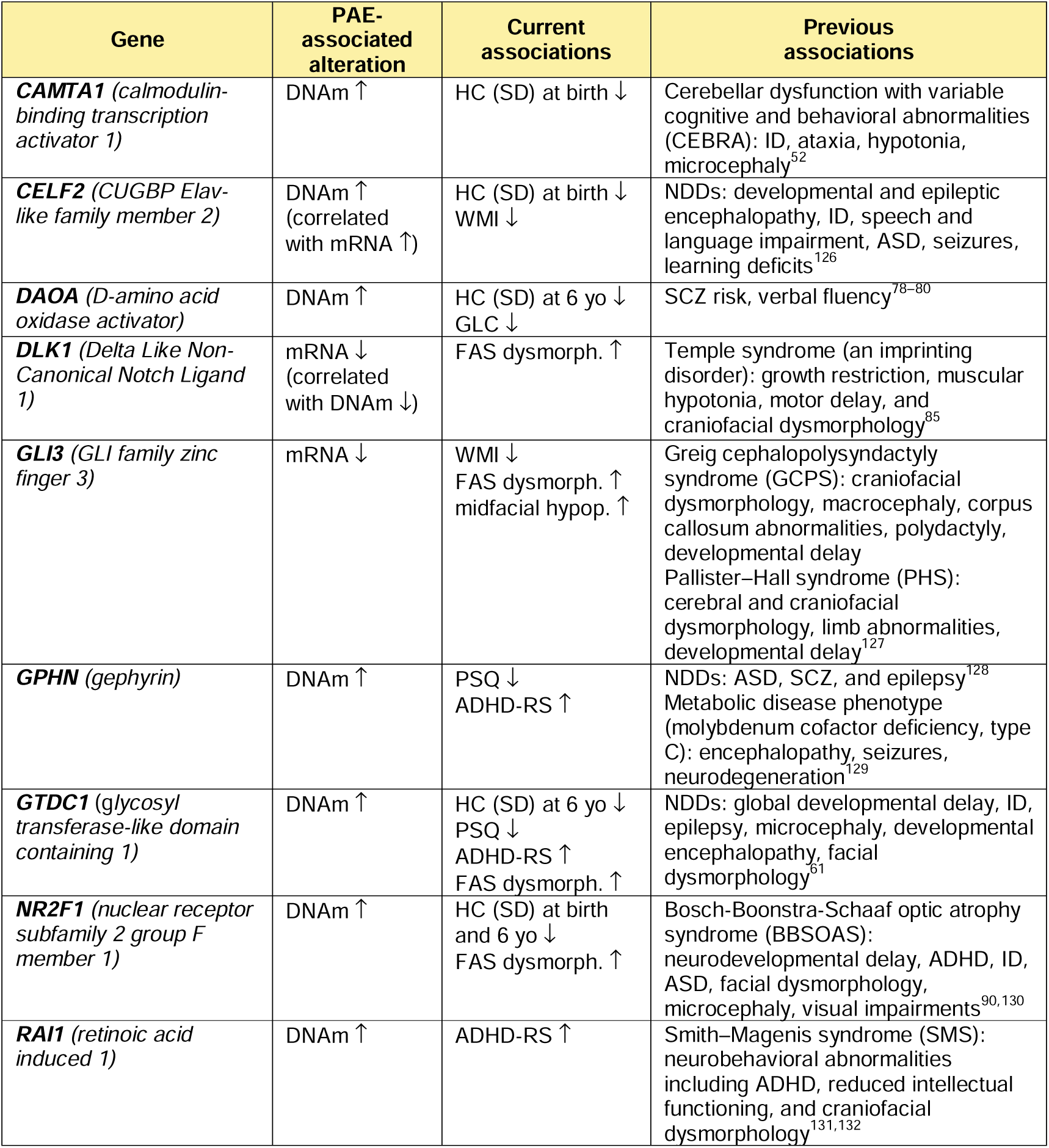
Selected genes associated with PAE and phenotypic features in the current and previous studies. The directions in the observed alterations in DNAm and gene expression (hypo ↓ /hyper ↑ or downregulated ↓ /upregulated ↑) and phenotypic traits (decreased ↓ /increased ↑ HC (SD) or scores) have been indicated by arrows. ID: intellectual disability.

One of the most interesting candidate gene was nuclear hormone receptor *NR2F1,* a TF whose mutations cause Bosch–Boonstra–Schaaf Optic Atrophy Syndrome (BBSOAS) including developmental delay, intellectual disability, visual impairment, epilepsy, autistic traits, craniofacial malformations as well as motor, behavioral, and brain structure abnormalities^63^. It has a crucial role in neuronal development and function, including neuronal proliferation and differentiation, development of neural crest, migration of cortical neurons, formation of neuronal circuits and connectivity, and neuronal E/I balance^63^. Interestingly, upregulated *NR2F1* expression has been observed in alcohol-exposed mouse embryonic cells differentiated towards neuroectoderm^133^, human cerebral organoids engrafted into ethanol-exposed mice, and PAE human fetuses^122^. In the current study, increased *NR2F1* DNAm was associated with decreased HC (SD) of newborns and six-year-olds, as well as increased FAS dysmorphology scores and maternal alcohol consumption (maternal AUDIT scores and alcohol dose of the full cohort).

Among the most interesting FASD candidate genes were hypermethylated *GPHN* and downregulated *GLI3*. *GPHN* is a scaffold protein considered as a central regulator of neuronal functions, which clusters GABA alpha receptor on the postsynaptic membrane at inhibitory GABAergic and glycinergic neurons^49^. It has been previously associated with several NDDs, such as ASD, SCZ, and epilepsy^128^ (Table 3). Furthermore, *GPHN* is involved in molybdenum cofactor biosynthesis and mutations in it cause a rare metabolic disease, Molybdenum cofactor deficiency type C with neonatal seizures and progressive encephalopathy^129^. In addition to placental hypermethylated *GPHN*, we observed upregulated *GABARAP*, which binds to *GPHN* and is involved in trafficking and localization of GABA alpha receptors^50^. These changes, which have no previous associations with PAE, may indicate altered clustering and function of GABA alpha receptors, disrupted inhibitory synaptic function, and remodeled neuronal networks. Interestingly, *GPHN* downregulation has been previously observed in the postmortem hippocampi from alcoholics and cocaine addicts^46^. Furthermore, altered development and reduced numbers of GABAergic interneurons have been reported in human fetal and infant brain with PAE^134^ as well as a selective reduction of cerebral cortex GABA neurons in a late-gestation FASD mouse model^135^. Reduced GABA levels have been linked to diminished cognitive control and ADHD^136–138^, both of which are characteristic features of the FASD phenotype^139^. Consistently, in the current study, the increased DNAm of *GPHN* was associated with decreased PSQ and increased ADHD-RS scores at the age of six. Also, association with maternal alcohol consumption (maternal AUDIT scores of the full cohort and tested six-year-olds) was observed.

*GLI3* is a TF that mediates the Sonic hedgehog signaling pathway, which is critical for brain and craniofacial development^140^. It is required for proper hippocampal growth and organization, a structure essential for spatial memory and learning, and its expression was downregulated by alcohol exposure during neurulation stage in a previous mouse study^81,107^. Consistent with the previous findings, placental decreased *GLI3* expression correlated with decreased WMI scores as well as increased FAS dysmorphology and midfacial hypoplasia scores of the six-year-olds in this study.

### Effects of PAE on developmental programming

Several PAE-associated genes identified in this study are involved in early embryonic patterning, including *GLI3, MNX1, NODAL,* and the imprinted gene *MEST*. Additional developmental candidates included *FRZB* and *DKK1*, antagonists of canonical Wnt signaling, which were associated with midfacial hypoplasia at age six. Considering the central role of Wnt signaling in neural crest development and craniofacial morphogenesis^141^, and its known disruption following PAE^101^, these findings are biologically plausible. We observed increased DNAm at a regulatory region of *FRZB*, which was associated with reduced *FRZB* expression and midfacial hypoplasia. As tightly regulated Wnt signaling is essential for midfacial development, and *FRZB* has previously been implicated in neural crest-derived craniofacial morphogenesis in zebrafish^142^, altered *FRZB* expression may contribute to the mechanisms underlying PAE-associated midfacial dysmorphology. Notably, this finding provides preliminary evidence linking PAE-associated molecular alterations in both DNAm and gene expression to a subsequent human phenotype. However, functional studies are needed to establish causality and determine how altered *FRZB* regulation affects craniofacial development.

Additionally, the increased DNAm at the regulatory region of *CELF2*, which encodes RNA-binding protein regulating alternative splicing and mRNA stability during neuronal development, correlated with increased *CELF2* expression. Furthermore, DNAm of two other *CELF2* probes associated with HC (SD) at birth and WMI at the age of six. Considering *CELF2*’s previously observed role in cortical maturation, synaptic development, as well as learning and memory^126^, altered regulation may contribute to broader developmental perturbations resulting in both structural and functional neurodevelopmental outcomes following PAE.

Furthermore, other interesting FASD candidate genes were *DLK1, LUZP1*, and *LAMC3*, of which placental downregulation correlated with decreased DNAm on regulatory region as well as associated with phenotypic features at the age of six. Placental decreased expression of imprinted *DLK1,* which has been previously associated with Temple syndrome with restricted growth and specific craniofacial phenotype^85^, correlated negatively with the FAS dysmorphology scores in the current study. Decreased expression of *LUZP1* (*leucine zipper protein 1*), a cytoskeletal and centrosome-associated protein involved in cell division, ciliogenesis, cell signaling, cranial neural tube development as well as learning and memory in mouse^143,144^, correlated here positively with WMI and negatively with ADHD and midfacial hypoplasia scores at the age of six. Furthermore, the downregulation of *LAMC3* (*laminin subunit gamma-3*), which has previously been associated with occipital cortical development^145^, correlated here with PIQ scores.

All of these observed and biologically intriguing connections need to be further explored to understand how PAE influences developmental programming and contributes to later-life complex phenotypic outcomes. Considering the high number of PAE-associated neurodevelopmental genes identified in our study, our findings are consistent with the well-established sensitivity of the developing nervous system to alcohol exposure during developmental programming^12,13,146^. Interestingly, dysregulation of E/I balance is associated with various NDDs such as ADHD^147^, ASD^148^, and SCZ^149^, and a large proportion of the PAE-associated genes observed in the current study have been previously associated with these disorders. Our results suggest that, despite their distinct origins, these NDDs share genes and networks, which may contribute to their partially overlapping phenotypes^3,4^. Furthermore, since several addiction-associated genes were identified in the PAE placentas, the developmental programming of addiction should also be explored in future studies.

### Placenta as a molecular window into the early development

Functional grouping of genes using AI and GO terms revealed that the largest proportion of placental genes which associated with child phenotypes was involved in transcriptional and epigenetic regulation. Furthermore, the majority of the altered placental DNAm was associated with genes involved in synaptic and neuronal function, neuronal development, and signaling in development. This is consistent with increasing number of studies, which suggest that epigenetic regulation plays a significant role in the etiology of NDDs^150^ and emphasize a connection between placenta and brain.

Our findings suggest that the extraembryonic placenta—the earliest organ to form—may act as a mirror, reflecting alcohol-induced changes in gene regulation during early development across all fetal tissues, including the brain. Since alcohol exposure has been associated with alterations in the methionine cycle and DNAm^12^, which play a key role in gene regulation, the role of the epigenome in alcohol-induced changes is of particular interest. The persistence of PAE-induced DNAm changes across cell divisions and across different cell types would be possible through *DNA methyltransferase 1*, which maintains mitotic epigenetic memory. This is supported by cross-tissue DNAm alterations in the same genes in previous PAE studies, including *TACR3* hypomethylation observed in both the placenta and newborn ectodermal BECs^23^. Importantly, if a gene is not actively transcribed in the studied tissue, DNAm changes do not necessarily result in altered gene expression. This may explain why many of the neuronal genes exhibiting altered DNAm in the placenta were not expressed in placental tissue, whereas the most pronounced expression changes were observed in genes involved in mitochondrial function and ribosomal translation, processes that are essential for placental physiology.

Although the molecular mechanisms remain poorly understood, the idea that the placenta could serve as a valuable tool for elucidating human brain development and associated disorders is appealing. This possibility deserves future exploration as it could revolutionize the understanding of complex developmental disorders. Moreover, the PAE-associated cross-tissue DNAm changes could provide valuable biomarkers for diagnosing disorders.

### Limitations

We are aware of the limitations in this study. Because the PAE-associated genes were identified in the placenta, it is unknown whether these findings extend to embryonic tissues, especially the brain. Moreover, the causality between the observed molecular alterations and neuropsychological performance at six years of age remains unknown. Furthermore, because we aimed to focus on alcohol-specific effects, genes influenced by both smoking and alcohol may not have been retained among the significant molecular changes and were therefore not included in the phenotype association analyses. Although the molecular effects of relatively high PAE were detectable in this cohort, the limited sample size precluded analyses of genomic influences, and potential contributions of other prenatal exposures and postnatal environmental factors cannot be ruled out. Finally, information on alcohol consumption was derived mainly from maternal self-reports collected in a specialized outpatient clinic for pregnant women with substance use issues, which may be subject to inaccuracy.

## CONCLUSION

In this study, by using an epigenetic approach and placental tissue, we bring forth PAE-induced alterations in DNAm and gene expression, that were associated with the neuropsychological and dysmorphological phenotypes of the same children at the age of six. In addition to identifying numerous candidate genes for the phenotypic features of FASD, this work highlights the placenta as a valuable tissue for understanding early human development and developmental disorders.

## MATERIALS AND METHODS

### The epiFASD cohort

Informed consent has been obtained from all participants, and we have an ethical approval (HUS/683/2020) as well as permission from Helsinki University Hospital (HUS/706/2025) for the epiFASD cohort. Women (*n*=87) with substantial alcohol consumption were recruited to this study in a special outpatient clinic for pregnant women with substance use problems in Helsinki University Hospital, Finland during the years 2013–2020 (Supplementary Table S1). All newborns were children of Finnish, Caucasian mothers. The amount and timing of maternal alcohol consumption was registered using self-reported information. To avoid specific individual level data, maternal alcohol consumption is presented in figures in three categories according to Alcohol Use Disorders Identification Test (AUDIT) scores or the number of alcohol units consumed per week (alcohol doses) (one unit is 12 g of ethyl alcohol in Finland)^151,152^. A 10-item screening tool AUDIT, developed by the World Health Organization, estimates alcohol consumption, drinking behavior, and alcohol-related problems. AUDIT scores 1–5 suggest low-risk consumption or <7 alcohol doses cause low risk for morbidity and mortality for non-pregnant women (category 1), scores 6–13 suggest hazardous or harmful alcohol consumption or 7–11 alcohol doses cause moderate risk for morbidity and mortality for non-pregnant women (category 2), and scores 14–40 indicate the likelihood of alcohol dependence (moderate-severe alcohol use disorder) or ≥12 alcohol doses cause high risk for morbidity and mortality for non-pregnant women (category 3) (Supplementary Fig. S2). The timing of consumption is also presented in figures in three categories according to the length of alcohol exposure: GW 1–12, GW 1–28, and GW 1–42. However, self-reported information (not categories) about timing of drinking, AUDIT scores, and alcohol doses were used in statistical analyses. 18 (20.7%) of the deliveries were cesarean sections (CS). According to the medical charts, the majority, 71 (81.6%) mothers of all PAE newborns smoked, and 23 (26.4%) mothers used antidepressants or antipsychotic medication during the pregnancy. Five (5.7%) mothers used gestational diabetes mellitus medication. Four mothers had thyroid diseases, three had antihypertensive medication, and one had preventive medication for herpes. One mother had an FAS diagnosis. Two mothers were occasional users of stimulants and four of cannabis. Due to the preterm premature rupture of membranes, two newborns were preterm. One of the PAE newborns had polydactyly, and three had a cleft lip.

The control samples (*n*=77), collected during the years 2013–2015 in Helsinki University Hospital, Finland, were from newborns of healthy Finnish, Caucasian mothers who did not use alcohol or tobacco during pregnancy according to their self-reported information (Supplementary Table S1). Five (6.5%) of the deliveries were CSs. The SEO samples (*n*=11), collected during the years 2013–2020 in Helsinki University Hospital, Finland, were from newborns of healthy Finnish, Caucasian mothers who smoked throughout pregnancy but did not use alcohol, according to their self-reported information (Supplementary Table S1). One (9.1%) of the deliveries was a CS.

Differences in newborn birth measures and maternal characteristics between study groups were calculated, depending on the data distribution, by Pearson’s chi-square test, parametric two-tailed Student’s t-test, or nonparametric Mann-Whitney U test. Differences in birth weight, birth length, and HC of the newborns were calculated using both anthropometric measures and the SDs of measures based on Finnish growth charts^153^, in which gestational age at birth, twinning, parity, and sex were considered when calculating the SDs (z-scores) of birth measures.

### Phenotypes of the epiFASD cohort children at the age of six

A total of 83 six-year-old children of the epiFASD cohort who were born in 2014-2018 participated in neuropsychological and dysmorphological assessments during 2020-2025: 30 alcohol-exposed, 48 unexposed controls, and five SEO. The following neuropsychological tests were used: WPPSI-III (WPPSI-III - Wechsler Preschool and Primary Scale of Intelligence - Third Edition) and WISC-IV (Wechsler Intelligence Scale For Children – IV). ADHD-RS (ADHD Rating Scale IV) questionnaires were filled out by parents and preschool/kindergarten teachers. Detailed dysmorphological assessments were performed. Minor anomalies and notable features were recorded and a FAS dysmorphology score was calculated based on an adaptation of Hoyme et al. (2016)^154^ with minor modifications. Detailed descriptions of the tests and tested children are provided in Jolma et al. (2026)^13^.

### Sample collection and preparation

Placental biopsies of newborns were collected immediately after delivery. When this was not possible, placenta was stored in the fridge for a maximum of 12 h and only DNA was extracted for further analyses. Biopsies (1 cm^3^) were collected from the fetal side of the placenta within a radius of 2–4 cm from the umbilical cord, rinsed in cold 1× PBS, and stored in RNAlater® (Thermo Fisher Scientific) at −80 °C.

Placental genomic DNA was extracted from one to four (3.2 on average) pieces of placental tissue samples using commercial QIAamp Fast DNA Tissue or AllPrep DNA/RNA/miRNA Universal Kits (Qiagen) or standard phenol-chloroform protocol. RNA was extracted from the same placental pieces as DNA (2.9 pieces on average) by AllPrep DNA/RNA/miRNA Universal Kit. RNA quality was assessed using an Agilent 2100 Bioanalyzer (Agilent Technologies, Inc.), which was provided by the Biomedicum Functional Genomics Unit (FuGU) at the Helsinki Institute of Life Science and Biocenter Finland at the University of Helsinki.

### DNAm microarrays

#### DNAm measurements and data processing

Genomic DNA (1000 ng) from available placental samples was sodium bisulfite-converted using the Zymo EZ DNAm™ kit (Zymo Research), and genome-wide DNAm was assessed with Infinium Methylation EPIC BeadChip Kit (Illumina) following a standard protocol at the Institute for Molecular Medicine Finland.

The raw DNAm dataset was pre-processed, quality controlled, and filtered with ChAMP R package using the minfi method^155^. Quality control was performed with the following settings: the detection *P*-value cutoff was set at *P*=0.01, probe bead count >3 in at least 95% of samples, and the threshold for failed probes’ ratio in each sample was set at 0.1. All samples passed these quality control thresholds, but 61 794 probes were excluded from the subsequent steps. Type-I and Type-II probes were normalized using the adjustedDasen method in the wateRmelon R package^156,157^. Potential effects caused by technical factors and biological covariates were studied from singular value decomposition (SVD) plots. The data was corrected for the effects of array, slide, analysis batch, and DNA extraction method using the Empirical Bayes method implemented in ComBat function in the R package sva^158^. After ComBat adjustment, probes located in sex chromosomes and probes binding to polymorphic and off-target sites were filtered out^159,160^. Also, probes in Finnish-specific single nucleotide polymorphisms (SNPs), which overlap with any known SNPs with global minor allele frequency, were removed. Population-specific masking information was obtained from Zhou et al. (2017)^161^.

Subsequently, a total of 616 912 probes were retained for further downstream analysis. Annotation information was merged to corresponding probes from the IlluminaHumanMethylationEPICanno.ilm10b4.hg19 R package^162^, which is based on the file “MethylationEPIC_v-1-0_B4.csv” from Illumina. Probe genomic locations relative to gene were annotated based on GENCODE Comprehensive V12 (GencodeComp) database. The following abbreviations were used: TSS1500: 1500 bp upstream of transcription start site (TSS), TSS200: 200 bp upstream of TSS, UTR: untranslated region, N_shelf: north shelf, N_shore: north shore, S_shore: south shore, S_shelf: south shelf. Probes were annotated to genes based on the University of California, Santa Cruz (UCSC) and GencodeComp databases.

The Planet R package^163^ was used to estimate placental cell-type fractions in each sample by CIBERSORT method in the epiDISH R package^164^. To calculate the differences in placental cell type composition, parametric two-tailed Student’s t-test or non-parametric Wilcoxon rank-sum exact tests were used. The statistical tests were selected based on assessments of data normality using the Shapiro–Wilk test and homogeneity of variances using the F-test. We observed no significant differences between the study groups in any placental cell-type.

#### Differentially methylated position analysis

Genome-wide differential DNAm analysis by using M-values was performed by the limma R package^165^, and the linear model was adjusted based on SVD plots for technical covariates batch and DNA extraction method, biological covariates newborn sex, maternal smoking, as well as cell-type fractions. β-values were used for visualization and interpretation of the results. DMPs were considered as significant when the DNAm difference in the effect size was ≥5% (Δβ ≤ −0.05 and Δβ ≥ 0.05) and the FDR-corrected *P*-value smaller than 0.05. Benjamini-Hochberg procedure was used to control for FDR.

#### Sensitivity analysis for DMPs

To test the sensitivity of DMPs, only samples without smoking exposure (16 PAE and 77 controls) were selected to the differential DNAm analysis, which was performed in limma and adjusted for newborn sex, batch, DNA extraction method, and cell-type fractions. Due to inflation, bacon R package was used to calculate inflation-adjusted CpGs^166^. CpGs with bacon-corrected FDR smaller than 0.05 were considered significant.

#### DMR analysis

The DMRcate R package was used for analysing DMRs^167^. DMRcate was set to determine probes (≥3) in a region with a maximum allowed genomic distance of 1000 bp, and scaling factor C set at 2, as per authors’ recommendation. Default *P*-value cut-offs, including FDR<0.05 for individually significant CpGs, were used in the analysis.

#### Pathway analyses

Enrichment analysis was performed for PAE-associated differentially methylated CpGs (FDR<0.05) by gometh function in the missMethyl R package^168^, which considers the different number of probes per gene present on the EPIC array and CpGs that are annotated to multiple genes. Enrichment analysis was also performed for PAE-associated DMRs by goregion function in missMethyl. The GO knowledgebase was used for identifying significantly enriched biological process (BP), cellular component (CC), and molecular function (MF) terms. Benjamini-Hochberg procedure was used to control for FDR.

#### Association analyses between PAE-associated probes and phenotypic features

Associations between DMPs and significant probes (FDR<0.05) associating with the same genes as DMPs, maternal alcohol consumption, HC (SD) at birth and at the age of six, cognitive performance, ADHD-RS scores, FAS dysmorphology scores, and midfacial hypoplasia were calculated with linear regression models in the CpGassoc R package^169^. Adjusted β-values were used as the dependent variable, phenotype as the independent variable, and the same covariates as in the main DMP analysis were included in these models. The number of samples included in the analyses varies depending on the availability of DNAm and phenotype data. For probes linked to multiple transcripts mapping to different chromosomes, possible off-target binding was assessed following a method from Chen et al. (2013)^170^.

### 3’mRNA sequencing (mRNA-seq) analysis

#### Differential expression analysis

Extracted RNA was prepared by diluting samples to 10 ng/μl and the library preparation for bulk 3′-sequencing of poly(A)-RNA was performed as previously described^171^ provided by FuGU. The libraries were sequenced on NextSeq 500 (Illumina). Drop-seq pipeline was used to construct the mRNA-seq count table for available placental RNA samples provided by FuGU. After filtering, a total of 20 851 transcripts were identified for downstream analysis. Principal component analysis implemented in the DESeq2 R package^172^ was used to identify batch effects, and ComBat-seq^173^ was used to adjust separate mRNA-seq batches. Differential expression analysis was performed by DESeq2, with model adjusting for smoking and newborn sex. DEGs were considered significantly differentially expressed when FDR-corrected *P*-value was <0.05. Benjamini-Hochberg procedure was used to control for FDR.

#### Sensitivity analysis for DEGs

To test the sensitivity of DEGs, only samples without smoking exposure (10 PAE and 41 controls) were selected to the differential expression analysis, which was performed in DESeq2 and adjusted for newborn sex.

#### Pathway analysis

enrichGO function in the clusterProfiler R package^174^ was used to perform enrichment analysis for DEGs (FDR<0.05). The GO knowledgebase was used for identifying significantly enriched BP, CC, and MF terms. Benjamini-Hochberg procedure was used to control for FDR.

#### Correlations between PAE-associated genes and phenotypic features

Correlations between DEGs, maternal alcohol consumption, HC (SD) at birth and at the age of six, cognitive performance, ADHD-RS scores, and FAS dysmorphology scores were calculated with Spearman’s rank correlation. Correlation between DEGs and midfacial hypoplasia was calculated with point-biserial correlation. The number of samples included in the analyses varies depending on the availability of gene expression and phenotype data. Vst expression counts were used. Bonferroni-adjusted significance thresholds were defined as 0.05 divided by the number of phenotypic variables multiplied by the number of DEGs. Analyses were performed in IBM SPSS Statistics, version 30.0 (IBM Corp.).

### Database comparisons

MitoXplorer 3.0 web tool^34^ was used to identify genes associated with mitochondrial processes. SynGO knowledgebase^38^ was used to investigate the associations to synapse function. Databases SZDB2.0^41^ for SCZ-associated genes and SFARI Gene^40^ for ASD-associated genes, as well as genes associated with ADHD according to GeneCard^39^ were chosen to assess common genes due to partly overlapping phenotypes with FASD.

### Correlation analysis between DNAm and gene expression

Pearson correlations were calculated between DNAm at the regulatory regions and gene expression in two ways: (1) correlation between DMPs and differentially methylated CpGs (FDR<0.05), which associated with the same genes as the DMPs, and corresponding gene expression, and (2) correlation between expression of DEGs and CpG sites in the corresponding gene. In the analyses, CpGs annotated exclusively for TSS1500, TSS200, 5′ UTR, and 1stExon regions, based on GencodeComp annotation, were considered as regulatory region CpGs. A total of 8692 genes expressed at a substantial level, defined as counts of 10 or above in at least 41 samples (number of samples in the control group) were included in the analysis. Adjusted β-values of CpGs and vst expression counts were used. Bonferroni correction was used for multiple testing correction.

### Functional groups

Functional clusters of PAE-associated HC, cognition, ADHD, and FAS dysmorphology genes were assembled based on GO terms with assistance of artificial intelligence (ChatGPT-4.1) and SynGO knowledgebase^38^. The identified clusters and associated genes were edited and verified by the authors.

### Artificial Intelligence

In addition to functional clustering of genes, language refinement was performed with assistance of artificial intelligence (ChatGPT-4.1, Microsoft Copilot).

## Data Availability

All data produced in the present work are contained in the manuscript.

## ACKNOWLEDGEMENTS

We express our sincere gratitude to all the families who participated in this study. We also thank PhD Heidi Marjonen-Lindblad, as well as research nurses Teija Karkkulainen and Jaana Palukka, for their invaluable contributions. The authors wish to acknowledge CSC – IT Center for Science, Finland, for computational resources.

