## Supplementary Figures for "Changes in neuronal genes in prenatally alcohol-exposed placentas associate with neuropsychological traits at the age of six"

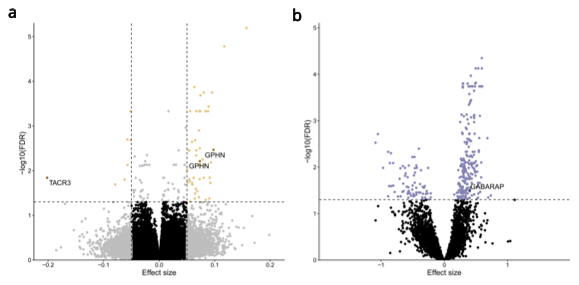

**Supplementary Figure S1.** PAE-associated differential DNAm and gene expression in the sensitivity analyses. **a)** Volcano plot showing the distribution of associations between placental CpG sites and PAE. Horizontal line marks FDR 0.05 and vertical line marks effect size ± 0.05. **b)** Volcano plot showing the distribution of associations between placental mRNA expression and PAE. Horizontal line marks FDR 0.05.

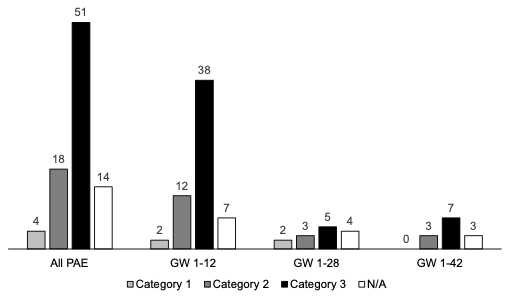

**Supplementary Figure S2.** Timing and amount of maternal alcohol consumption in categories. Visual presentation of the timing and amount of alcohol consumption of the PAE mothers (n = 87) during gestational weeks (GW) 1–12, GW 1–28, and GW 1–42. Categories for maternal alcohol consumption: AUDIT scores 1–5 suggest low-risk consumption or <7 alcohol units consumed per week (alcohol doses) cause low risk for morbidity and mortality for non-pregnant women (category 1), AUDIT scores 6–13 suggest hazardous or harmful alcohol consumption or 7–11 alcohol doses cause moderate risk for morbidity and mortality for non-pregnant women (category 2), and AUDIT scores 14–40 indicate the likelihood of alcohol dependence (moderate-severe alcohol use disorder) or ≥12 alcohol doses cause high risk for morbidity and mortality for non-pregnant women (category 3). The categories for maternal alcohol consumption were defined by combining information from previous publications (Rehm et al. 2015, Tynjälä et al. 2012).
